# Persistent *Borrelia burgdorferi* protein antigen in CD16^+^ monocytes is associated with monocyte-skewed immune dysregulation in post-treatment Lyme disease syndrome (PTLDS)

**DOI:** 10.64898/2026.09.24.26363601

**Authors:** Bruce K. Patterson, Edgar B. Francisco, Nicolas Castaño, Kwat Medetgul-Ernar, Christopher Beaty, Kaeden Beaty, Patricia A. Felder, Ram Yogendra

**Author notes:** Address correspondence to Bruce K. Patterson, MD 10850 E. Traverse Hwy Suite 4400 Traverse City, MI 49684.

## Abstract

Post-treatment Lyme disease syndrome (PTLDS) is characterized by persistent symptoms after antibiotic therapy for *Borrelia burgdorferi* infection. The mechanisms underlying these symptoms remain incompletely understood and have been attributed to ongoing low-level infection, autoimmunity, or immune dysregulation driven by residual bacterial antigens. We investigated whether *Borrelia burgdorferi* protein antigen persists in long-lived monocyte subsets in PTLDS patients and is associated with broader immune dysregulation. CD14^⁺^CD16^⁺^ monocytes from 10 patients with PTLDS were analyzed by mass spectrometry; plasma from all 10 was profiled on a 14-plex cytokine panel against 50 healthy donors; PBMCs from 6 patients (3 antigen-positive and 3 antigen-negative) underwent RT-qPCR and bulk RNA-seq with immune-compartment profiling. FlaB and OspC were detected in CD14^⁺^CD16^⁺^ monocytes months to years after treatment, whereas *Borrelia burgdorferi* transcript was not detected. Plasma IFN-γ exceeded the healthy donor median in all 10 participants. Antigen-positive samples showed an antigen-associated inflammatory transcriptional profile, including increased TNFSF14/LIGHT, GSDMD, GBP5, and KCNJ2 expression, together with a compartment-level pattern of monocyte-skewed myeloid reconfiguration, higher neutrophil-related features, and selective T-cell-state changes. These findings link residual *Borrelia burgdorferi* protein in CD14^⁺^CD16^⁺^ monocytes to monocyte-skewed immune remodeling in PTLDS, paralleling observations in long COVID and supporting a model in which post-infectious syndromes are sustained by antigen-fueled immune remodeling rather than ongoing infection.

**Importance:** Many people treated for Lyme disease go on to experience fatigue, pain, and difficulty thinking for months or years, and the reason is unknown. Persistent infection has been a leading explanation, yet longer courses of antibiotics do not help. We found proteins from *Borrelia burgdorferi*, the bacterium that causes Lyme disease, inside long-lived blood monocytes of patients with lasting symptoms, years after treatment and with no sign that the bacterium was still active. Patients carrying these proteins showed a distinct inflammatory immune profile. These findings point to leftover bacterial material, rather than ongoing infection, as a possible driver of persistent symptoms, echo observations in long COVID, and argue that treatments directed at the immune response deserve study.

## Introduction

Persistent symptoms following resolution of acute infections, including Lyme disease and long COVID, represent a growing public health challenge. Lyme disease, caused by the spirochete *Borrelia burgdorferi*, is the most common vector-borne illness in North America, with approximately 476,000 cases diagnosed and treated annually in the United States (1, 2).

Transmitted primarily by Ixodes ticks, *Borrelia burgdorferi* disseminates through the bloodstream and connective tissues, leading to a range of clinical manifestations (2–4). Although most patients recover fully with timely antibiotic treatment, approximately 10–20% experience lingering symptoms such as fatigue, musculoskeletal pain, and cognitive impairment (5, 6). This condition, known as post-treatment Lyme disease syndrome (PTLDS), can significantly impair quality of life and remains incompletely understood. Proposed mechanisms include autoimmune responses, immune dysregulation, or prolonged inflammation triggered by residual bacterial antigens (6). The expanding geographic range of ticks and the complex course of Lyme disease highlight the need for continued research into its pathogenesis and sequelae.

If untreated, Lyme disease typically progresses through three stages. Early localized infection often presents with erythema migrans and flu-like symptoms such as fever, headache, and fatigue (4). In the early disseminated stage, spirochete spread can result in neurological complications or cardiac abnormalities, while late disseminated disease, occurring months or years after the initial infection, is characterized by chronic arthritis or neurocognitive impairment (4). Diagnosis is challenging, as erythema migrans is absent in up to 30% of cases (2), and serological assays often yield false negatives in early infection or false positives due to cross-reactivity (2). Although antibiotics such as doxycycline, amoxicillin, or cefuroxime are generally effective (4), a subset of patients develop persistent symptoms despite appropriate treatment.

*Borrelia burgdorferi* possesses several features that enable immune evasion and prolonged host colonization. Periplasmic flagella permits corkscrew motility, facilitating tissue invasion (3). The organism’s ability to evade immune clearance – through antigenic variation, complement inhibition, and biofilm-like structures – complicates eradication and may contribute to long-term immunologic consequences (6). These adaptations differentiate *Borrelia burgdorferi* from many bacterial pathogens and complicate treatment outcomes.

Recent murine work suggests that *Borrelia burgdorferi* peptidoglycan (PG)—a structural polymer of the bacterial cell wall—can persist in tissues after infection and may be sufficient to drive prolonged inflammatory responses even after apparent microbial clearance (7). In humans, *Borrelia burgdorferi* PG has been detected as a persistent antigen in post-antibiotic Lyme arthritis (e.g., synovial fluid), supporting antigen persistence in at least some clinical settings (8). However, it remains unproven whether persistent Borrelia antigens contribute to the systemic symptom complex of PTLDS (9).

Evidence from innate immune activation studies supports the plausibility of antigen persistence–driven inflammation, in which non-replicating *Borrelia burgdorferi* debris (including PG) remains immunostimulatory after bacterial killing/clearance. Salazar et al. demonstrated that live *Borrelia burgdorferi* elicited a significantly stronger transcriptional and cytokine response in human monocytes and peripheral blood mononuclear cells (PBMC) than did equivalent amounts of bacterial lysates (10). Notably, live spirochetes triggered robust secretion of TNF-α, IL-6, IL-1β, and IL-18, as well as IFN-β and interferon-stimulated genes—responses that were only partially TLR2-dependent and could not be replicated by synthetic TLR2 agonists or lysed bacteria. Heat-inactivated *Borrelia burgdorferi* was also sufficient to induce cytokine release, indicating that bacterial viability and replication are not required for innate immune activation. These findings suggest that phagocytosed, non-replicative *Borrelia burgdorferi* components can remain immunostimulatory within host cells.

A similar mechanism of non-replicative antigen persistence has been observed in long COVID (also termed post-acute sequelae of COVID-19, PASC) and post-COVID vaccine syndrome. In these syndromes, the SARS-CoV-2 spike protein, particularly the S1 subunit, has been detected in non-classical monocytes (NCM) months after infection or vaccination, even in the absence of viral replication (11). NCM are a longer-lived, endothelial-patrolling subset, making them a plausible reservoir for retained immunostimulatory antigen (12). These retained viral antigens are thought to contribute to persistent inflammation and symptomatology, further supporting the concept that long-term immunologic sequelae can arise from the retention of pathogen-derived material within the host.

In this study, we investigated whether *Borrelia burgdorferi*–derived proteins persist in human CD16⁺ monocyte subsets following infection, using sorted CD14⁺CD16⁺ monocytes as the proteomic target population, and whether antigen persistence was accompanied by changes in plasma cytokine concentrations, PBMC gene expression, and compartment-level immune activity. This population overlaps with the biology of CD16⁺ intermediate monocytes (IM) and NCM implicated in persistent SARS-CoV-2 antigen carriage. Our findings provide new insights into the potential role of bacterial antigen persistence, rather than ongoing infection, in the pathogenesis of PTLDS.

## Results

### Plasma cytokine/chemokine signature of PTLDS individuals

To characterize the systemic inflammatory context in which monocyte antigen persistence was assessed, plasma from all 10 PTLDS participants was profiled by multiplex bead-based assay and compared with values from 50 healthy donors (Figure 1). Concentrations spanned more than three orders of magnitude, and the analytes differed in how consistently they were raised. Relative to the healthy donor median, IFN-γ was elevated in all 10 participants (P = 0.002, exact sign test), IL-4 and VEGF in 9 of 10 (P = 0.022 for both), and IL-6 in 8 of 10 (P = 0.11). TNF-α was the exception, exceeding the healthy donor median in only 3 of 10 participants (P = 0.34). Only IFN-γ survived Bonferroni correction across the five analytes (threshold P = 0.010). Stratifying by monocyte antigen status showed a consistent directional pattern that the cohort size could not resolve. Median concentrations were higher in the three antigen-positive than in the seven antigen-negative participants for IL-6 (32.0 versus 3.2 pg/mL), IFN-γ (16.1 versus 2.9), TNF-α (14.1 versus 3.9) and VEGF (112.4 versus 34.4), but not for IL-4 (7.4 versus 13.2). No difference reached significance by exact Mann–Whitney testing (TNF-α P = 0.067, IL-6 P = 0.12, IFN-γ P = 0.24, VEGF P = 0.38, IL-4 P = 0.67), an expected result given that a three-versus-seven comparison cannot yield a two-sided P below 0.017 under any configuration of the data (Figure 1). Participant D7 (Table 1) had the highest IL-4 and VEGF concentrations in the cohort but was antigen-negative under the prespecified criterion requiring both FlaB and OspC, although FlaB alone was detected at 31% sequence coverage in that sample (Table 2). These measurements were cross-sectional, taken at a single timepoint months to years after antibiotic treatment. Together they indicate that inflammatory and vascular signaling remained detectable long after treatment, with a non-significant trend toward higher concentrations in participants carrying monocyte-associated *Borrelia burgdorferi* antigen.

**Figure 1.**
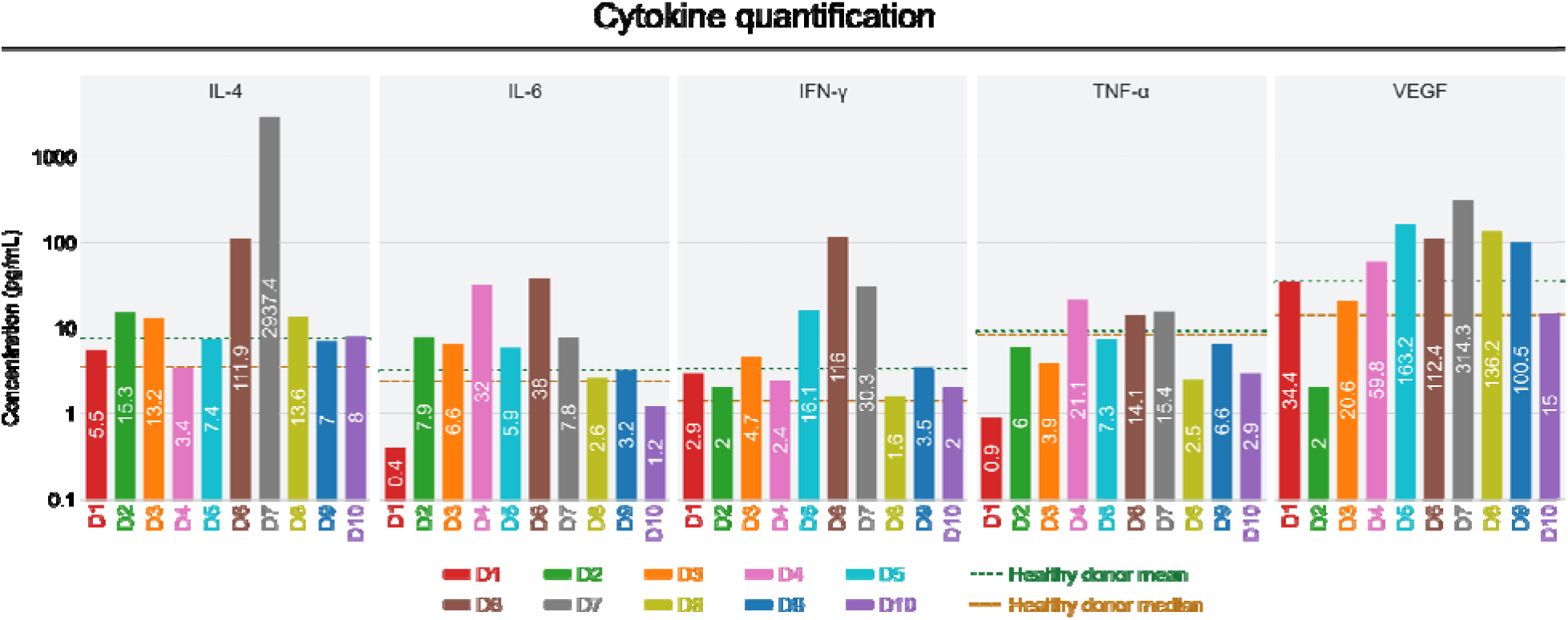
Plasma cytokine, chemokine, and growth factor concentrations in 10 PTLDS participants. Plasma collected at enrollment was assayed by a 14-plex bead-based flow cytometric assay. Five of the 14 analytes assayed are shown (IL-4, IL-6, IFN-γ, TNF-α, and VEGF). Each bar is one participant, labeled by donor ID as in Table 1, with values in pg/mL printed within each bar and plotted on a log₁₀ axis. Donors D4, D5, and D6 (P1–P3) carried detectable *Borrelia burgdorferi* FlaB and OspC in sorted CD14⁺CD16⁺ monocytes. The remaining seven were antigen-negative (Table 2). The dotted green line marks the healthy donor mean and the dashed orange line the healthy donor median for each analyte (healthy donor cohort: N = 50, assayed on the same platform and analyte panel). The healthy donor reference supports comparison against the healthy median rather than a two-sample test.

**Figure 2.**
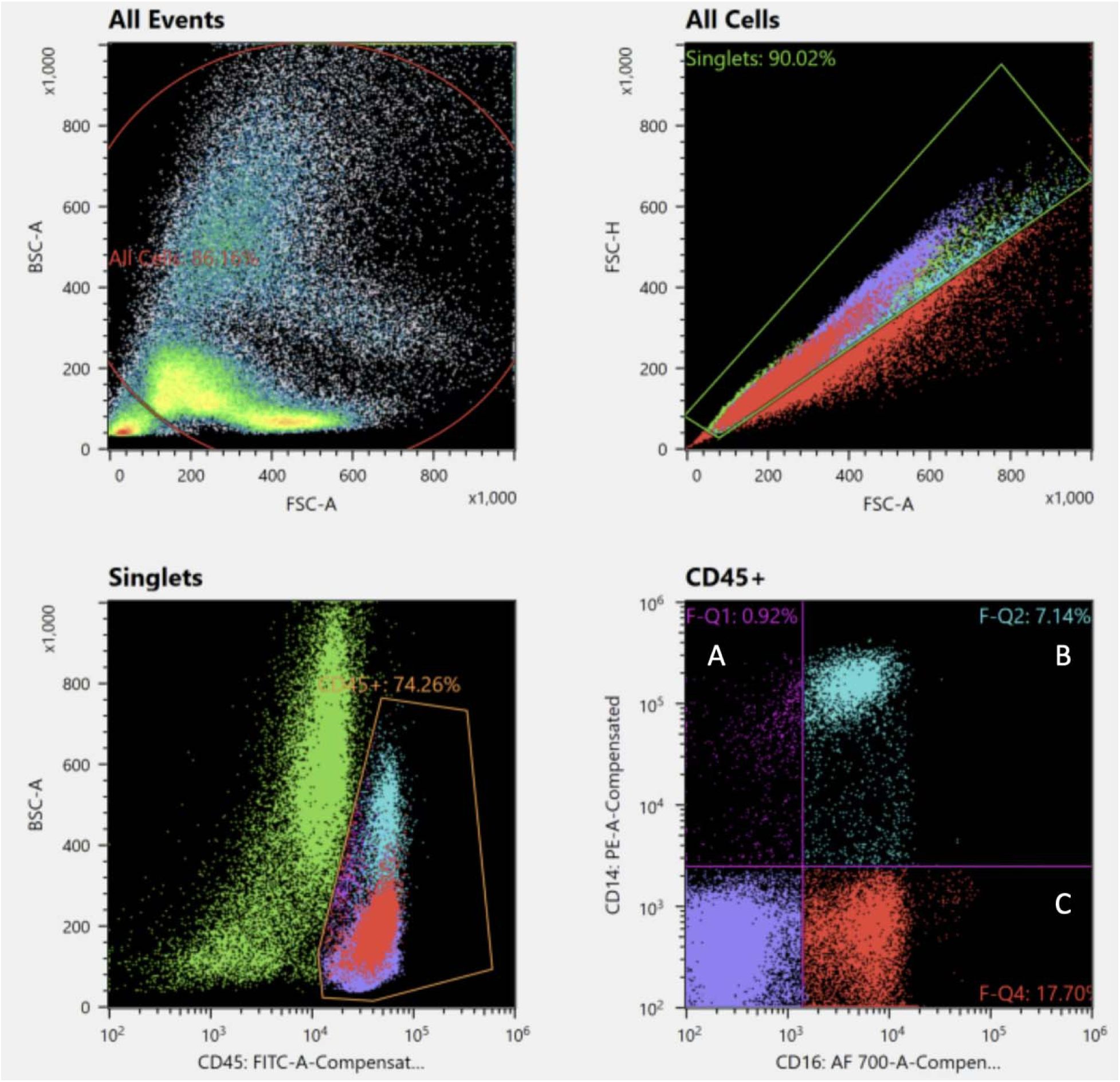
Representative flow sorting strategy for CD45⁺CD14⁺CD16⁺ monocytes. Events were first gated by FSC-A versus SSC-A to exclude debris, followed by singlet selection using FSC-H versus FSC-A. CD45⁺ leukocytes were then selected, and CD14 versus CD16 expression was used to identify monocyte subsets. The sorted proteomics population corresponded to CD45⁺ cells staining positive for both CD14 and CD16. This operational CD14⁺CD16⁺ gate captures CD16⁺ monocytes and was not further subdivided into intermediate and non-classical subsets for proteomic analysis.

**Table 1.** Patient demographics. Donors whose PBMCs were used for RNA-seq are indicated in parentheses. N1–N3 and P1–P3 denote PTLDS donors without and with, respectively, detectable *Borrelia burgdorferi* protein in sorted CD14⁺CD16⁺ monocytes.

| Donor | Sex | Age range (years) | Diagnosis (year) |
| --- | --- | --- | --- |
| D1 (N1) | Female | 51-55 | 2012 |
| D2 (N2) | Male | 16-20 | 2014 |
| D3 (N3) | Male | 16-20 | 2014 |
| D4 (P1) | Female | 61-65 | 2003 |
| D5 (P2) | Male | 41-45 | 2019 |
| D6 (P3) | Male | 26-30 | 2015 |
| D7 | Female | 56-60 | 2023 |
| D8 | Male | 66-70 | 2025 |
| D9 | Male | 76-80 | 2018 |
| D10 | Female | 46-50 | 2012 |

**Table 2.** A sample was classified as *Borrelia burgdorferi* antigen-positive only when both Flagellar filament core protein (FlaB) and Outer Surface protein C (OspC) were detected by mass spectrometry. N1–N3 and P1–P3 denote donors whose peripheral blood mononuclear cells (PBMC) were used for RNA sequencing. ND: not detected.

| Donor ID | Protein | Coverage (%) | Number of peptide spectrum matches | Number of peptides | Number of unique peptides | Classification |
| --- | --- | --- | --- | --- | --- | --- |
| D1 (N1) | FlaB | 11 | 6 | 3 | 3 | Negative |
|  | OspC | ND | ND | ND | ND |  |
| D2 (N2) | FlaB | ND | ND | ND | ND | Negative |
|  | OspC | ND | ND | ND | ND |  |
| D3 (N3) | FlaB | ND | ND | ND | ND | Negative |
|  | OspC | ND | ND | ND | ND |  |
| D4 (P1) | FlaB | 48 | 418 | 13 | 13 | Positive |
|  | OspC | 26 | 55 | 8 | 8 |  |
| D5 (P2) | FlaB | 36 | 40 | 7 | 7 | Positive |
|  | OspC | 17 | 17 | 3 | 3 |  |
| D6 (P3) | FlaB | 44 | 402 | 12 | 12 | Positive |
|  | OspC | 52 | 135 | 9 | 9 |  |
| D7 | FlaB | 31 | 41 | 8 | 8 | Negative |
|  | OspC | ND | ND | ND | ND |  |
| D8 | FlaB | 18 | 5 | 2 | 1 | Negative |
|  | OspC | ND | ND | ND | ND |  |
| D9 | FlaB | ND | ND | ND | ND | Negative |
|  | OspC | ND | ND | ND | ND |  |
| D10 | FlaB | ND | ND | ND | ND | Negative |
|  | Ock | ND | ND | ND | ND |  |

### *Borrelia burgdorferi* FlaB and OspC persist in CD14^+^CD16^+^ monocytes

Data-independent acquisition (DIA) mass spectrometry detected two *Borrelia burgdorferi* proteins in sorted CD14^+^CD16^+^ monocytes from a subset of patients (Table 2): flagellar filament 41 kDa core protein (FlaB) and outer surface protein C (OspC). Neither protein was detected in the negative control, and both were recovered from their respective protein standards (Table S1). Representative peptide-mapping plots demonstrated sequence coverage for both proteins across antigen-positive samples (Figure 3), derived from peptide identifications supported by representative fragment ion spectra (Figure S1). FlaB was detected with 48%, 36%, and 44% sequence coverage in samples D4, D5, and D6, respectively, whereas OspC was detected with 26%, 17%, and 52% coverage in the same samples. Quantitative peptide-spectrum match and peptide counts were highest in P1 and P3 and lower, but still supportive of detection, in P2 (Table 2). FlaB was detected without OspC in one of the three antigen-negative donors whose PBMCs were sequenced (N1, 11% coverage, 3 unique peptides), and in two antigen-negative donors in the cohort (D8, 18%; D7, 31%). Because classification required detection of both proteins, all were designated antigen-negative.

**Figure 3.**
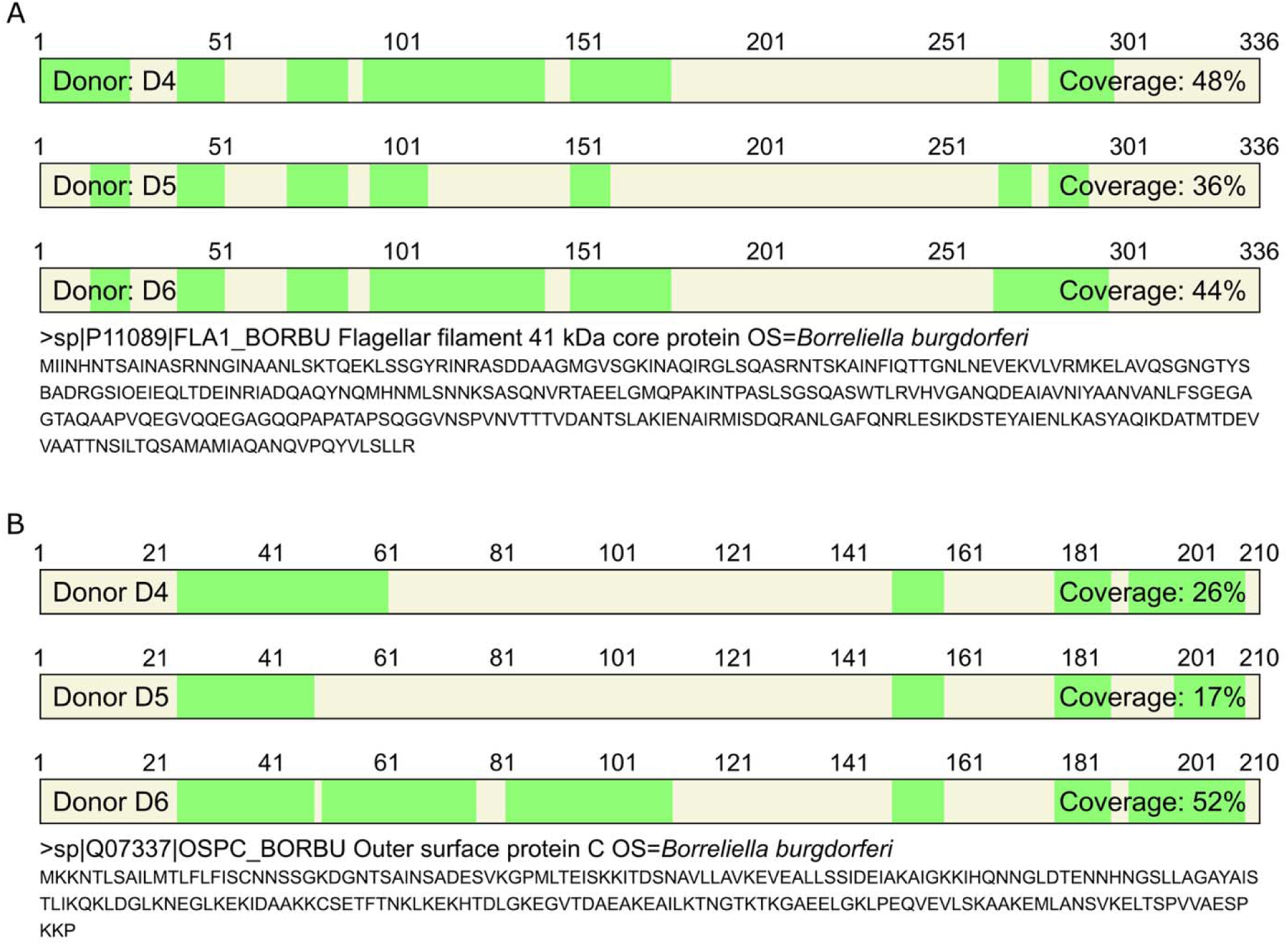
Representative protein sequence coverage of FlaB (A) and OspC (B) identified by proteomic analysis of sorted CD14⁺CD16⁺ monocytes from donors D4, D5, and D6. Peptides mapped to the protein sequences are highlighted in green.

### Lack of transcriptomic evidence for active *Borrelia burgdorferi* infection

Despite detection of *Borrelia burgdorferi*-derived proteins by mass spectrometry, RT-qPCR did not detect FlaB transcript in any of the six RNA-seq samples (Table S2). Appropriate amplification of the FlaB standard dilution series and RNase P amplification across all study samples confirmed assay performance, RNA integrity, and successful reverse transcription (Figure 4). Consistent with this, reads assigned to the *Borrelia burgdorferi* genome mapped exclusively to structural RNA loci — ribosomal RNA, transfer RNA, and tmRNA — with no protein-coding gene detected (Figure S2). Together, these findings support persistence of *Borrelia burgdorferi*-derived protein antigen without transcriptomic evidence of active infection. No *Borrelia burgdorferi* gene was differentially expressed between antigen-positive and antigen-negative samples after multiple-testing correction (P_adj_ > 0.25; Figure S2).

**Figure 4.**
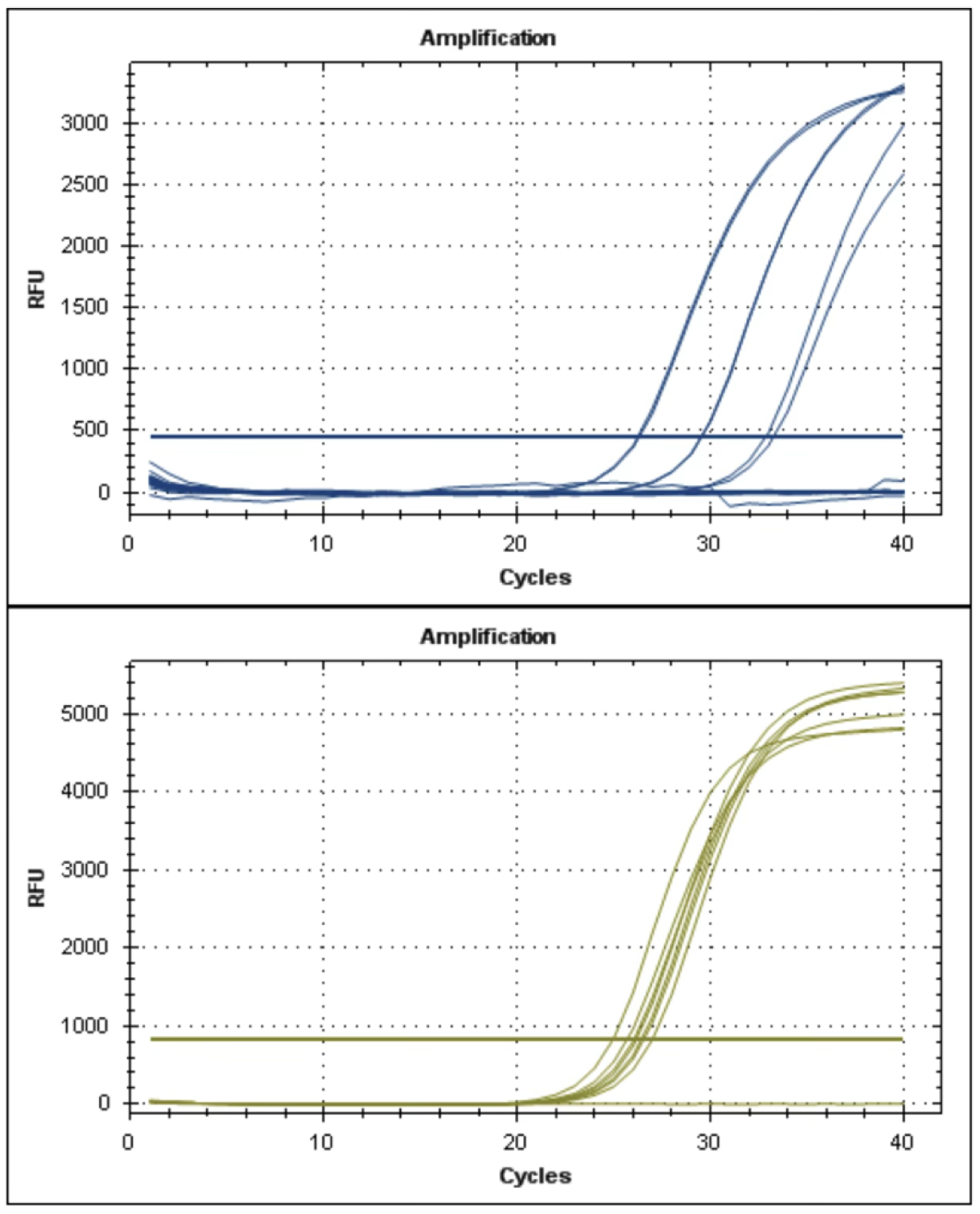
Representative amplification curves. (Top) *Borrelia burgdorferi* FlaB standard dilution series ranging from 10² to 10 copies. (Bottom) Human RNase P amplification across study samples, confirming amplifiable host nucleic acid and successful reverse transcription.

### Transcriptomic signature of individuals with antigen-bearing monocytes

Hierarchical clustering of the 65 genes differentially expressed at P_adj_ < 0.01 separated the two groups (Figure 5A). Across blood-expressed genes, 213 were differentially expressed at P_adj_ < 0.05: 144 higher and 69 lower in antigen-positive individuals (Figure 5B, Table S3). Principal component analysis (PCA) separated antigen-negative from antigen-positive samples along PC2, with the negative group clustering and the positive group more dispersed (Figure 5C).

**Figure 5.**
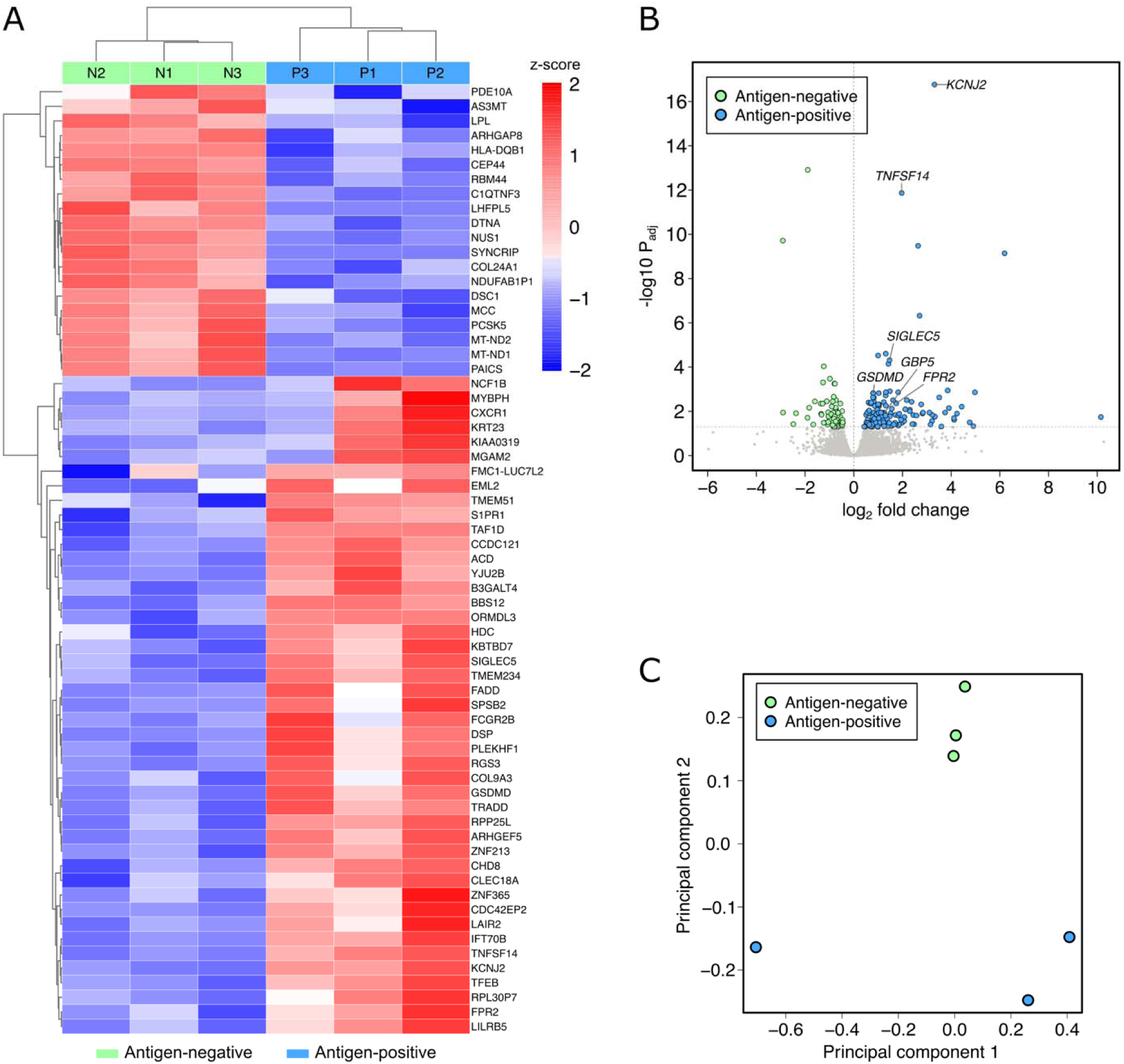
PBMC transcriptomes separated by monocyte *Borrelia burgdorferi* antigen status. PBMC RNA-seq from three donors without (N1–N3) and three with (P1–P3) detectable *Borrelia burgdorferi* protein in sorted CD14⁺CD16⁺ monocytes. (A) The 65 genes differing at P_adj_ < 0.01, clustered hierarchically. Cells are per-gene z-scores of log normalized counts, and the sample clustering recovers both donor groups. (B) All tested genes by fold change and significance, with the 213 genes differing at P_adj_ < 0.05 colored by direction. Positive log₂ fold change indicates higher expression in antigen-positive donors, and selected genes are discussed in the text. (C) Donors projected onto the first two principal components of variance-stabilized counts across the 500 most variable genes. Differential expression used PyDESeq2 with a Wald test and Benjamini–Hochberg adjustment, N = 3 per group.

Retention of *Borrelia burgdorferi* FlaB and OspC in monocytes was associated with a distinct transcriptional state that separated antigen-negative from antigen-positive donors in both hierarchical clustering and principal component analysis (Figure 5A,C). This antigen-positive signature was characterized by induction of inflammatory genes, including TNFSF14/LIGHT (log₂FC = 1.97, P_adj_ = 1.4 × 10⁻¹²), together with increased expression of GSDMD (log₂FC = 0.80, P_adj_ = 1.5 × 10⁻³), GBP5 (log₂FC = 1.21, P_adj_ = 0.013), IFIT2 (log₂FC = 1.72, P_adj_ = 0.038), and IFIT3 (log₂FC = 1.25, P_adj_ = 0.040), consistent with inflammasome and interferon signaling.

Additional upregulated genes included CXCR1 (log₂FC = 3.51, P_adj_ = 2.4 × 10⁻³), SIGLEC5 (log₂FC = 1.47, P_adj_ = 4.9 × 10⁻), FPR2 (log₂FC = 1.74, P_adj_ = 4.4 × 10⁻³), FCGR2B (log₂FC = 1.23, P_adj_ = 3.8 × 10⁻³), and LILRB5 (log₂FC = 1.32, P_adj_ = 5.6 × 10⁻³), indicating broader remodeling of chemotactic and immune receptor pathways. The most statistically significant gene was KCNJ2 (log₂FC = 3.32, P_adj_ = 1.7 × 10⁻¹). These data define an antigen-associated inflammatory transcriptional profile marked by TNFSF14 induction, and interferon- and inflammasome-associated transcripts.

Genes higher in antigen-positive donors were strongly enriched for transcripts characteristic of neutrophils in the healthy sorted immune-cell reference (60 of 144 genes, 41.7%, versus 8.6% of blood-expressed genes, odds ratio 7.95, P = 4.5 × 10⁻², Fisher’s exact test, Table S3), whereas the 69 genes higher in antigen-negative donors showed no such enrichment. The enrichment was carried by receptor and signaling transcripts, including CXCR1, FPR2, SIGLEC5, NCF1, NCF1B, FCGR2B and LILRB5. The enrichment could arise from differing granulocyte content between the groups rather than from monocyte gene expression, but no single granulocyte fraction accounts for the data. Reproducing the observed CXCR1 increase would require granulocytes to comprise approximately 14% of the PBMC pool, which predicts a 7.5-fold rise in CD177 and 1.9-fold rises in S100A8 and PADI4. Observed changes were 0.75-, 0.94- and 1.03-fold, with 95% confidence intervals excluding each predicted value. The dissociation between receptor and granulocyte-identity transcripts is therefore quantitative rather than a failure to detect change.

### Immunogram reveals monocyte-centric immune dysregulation

To determine whether residual *Borrelia burgdorferi* protein detection was associated with broader immune remodeling, we applied transcriptome-derived immune compartment profiling to the same six PBMC RNA-seq samples (Figure 6, Figure S3). Compared with the antigen-negative samples, antigen-positive samples showed higher general monocyte scores at the system level (Figure 6A) and higher IM/NCM and chronically activated macrophage scores in the monocyte sub-compartment (Figure 6B). Antigen-positive samples also had elevated neutrophil and low-density neutrophil signatures (Figure S3). By contrast, antigen-negative samples retained higher progenitor, naive T-cell, regulatory CD4 T-cell, naive CD8 T-cell, and naive B-cell scores (Figure 6A, Figure S3). Together, the antigen-negative profile retained features of intact progenitor output and naive replenishment across T and B lineages.

**Figure 6.**
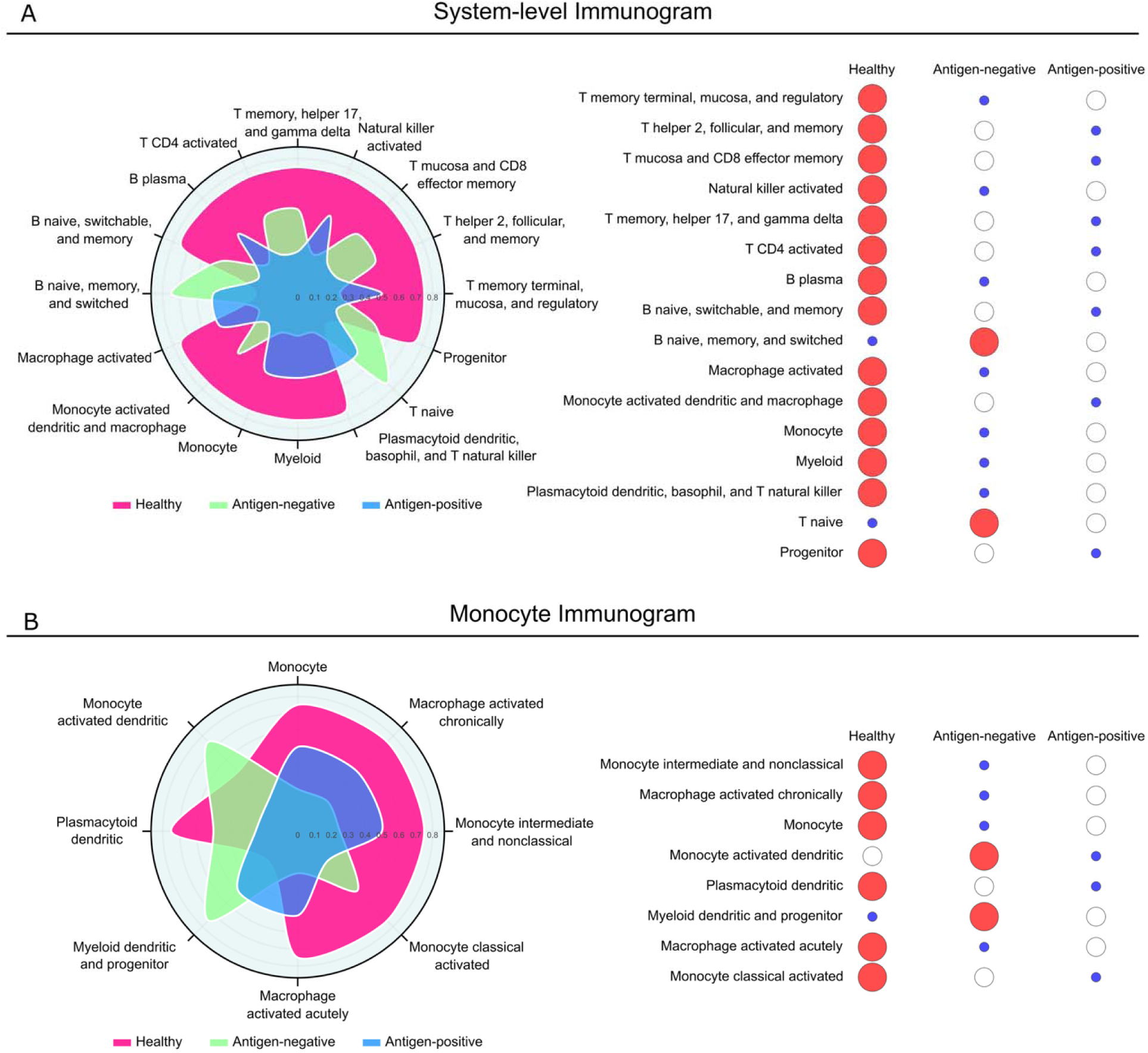
Immunogram profiles distinguish antigen-negative from antigen-positive PTLDS patients. (A) System-level Immunogram showing 16 cross-compartment immune activity axes. Antigen-positive samples show elevated monocyte and chronically activated macrophage axes together with reduced progenitor and naive T cell axes relative to antigen-negative samples. (B) Monocyte Immunogram resolving individual cell-state nodes within the monocyte and dendritic cell lineage. Antigen-positive samples show higher general monocyte and chronically activated macrophage scores together with reduced monocyte-activated dendritic cell, plasmacytoid dendritic, and myeloid dendritic/progenitor scores. Each panel shows a radar plot (left) and a bubble plot (right) of the same per-axis activity scores on a 0–1 scale. Antigen-negative samples are shown in green, antigen-positive samples in blue, and an external healthy reference cohort (GSE107011, N = 13) in magenta. The healthy cohort is shown for context only and was not included in statistical comparisons.

The largest contrast was within the monocyte compartment itself (Figure 6B). Although general monocyte and chronically activated macrophage scores were higher in antigen-positive samples, monocyte-activated dendritic, plasmacytoid dendritic, and myeloid dendritic/progenitor scores were reduced. Antigen-positive samples were therefore not uniformly enriched for all myeloid programs but instead showed a shift away from dendritic-cell-associated antigen-presenting differentiation.

Adaptive compartments also diverged by antigen status (Figure S3). Antigen-positive samples showed higher CD8 terminal memory scores and an elevated activated gamma-delta subnode within the broader T invariant compartment, while antigen-negative samples retained higher naive and regulatory T-cell scores. Within the B-cell compartment, antigen-positive samples showed elevated plasmablast scores alongside reduced plasma cell, progenitor, and naive/switchable B-cell scores, indicating B-cell activation without a corresponding expansion of terminally differentiated plasma cells or of the naive pool that would replenish it. Collectively, these findings indicate that residual *Borrelia burgdorferi* protein detection is associated with a monocyte-skewed inflammatory state, reduced dendritic-cell-associated antigen-presenting programs, and features of chronic adaptive immune stimulation.

The external healthy reference (GSE107011) showed higher Immunogram activity than both patient groups at nearly all nodes (54 of 65), placing both PTLDS groups below the healthy reference across most of the immune network (Figure 6, Figure S3). The two patient groups differed in which compartments were affected. Activity fell from the healthy reference to antigen-negative to antigen-positive participants across progenitor, naive, and regulatory nodes, and rose in that same order across myeloid and inflammatory nodes. Antigen status therefore tracked a redistribution of immune activity from renewal toward inflammation rather than a uniform loss of immune function.

## Discussion

*Borrelia burgdorferi* FlaB and OspC were detected in CD14⁺CD16⁺ monocytes from a subset of patients with PTLDS months to years after antibiotic treatment. In contrast, neither targeted FlaB RT-qPCR nor bulk RNA sequencing provided evidence of transcriptionally active Borrelia in these samples. Together, these findings support a model in which persistent bacterial antigen, rather than readily detectable active infection, is associated with chronic inflammatory signaling in a subset of PTLDS patients.

Persistent symptoms in PTLDS have been attributed to several non-mutually exclusive mechanisms. Proposed explanations include ongoing low-level infection or biofilm-like structures that evade antibiotics (6, 13), autoimmunity triggered by molecular mimicry between Borrelia antigens and host tissues (14), and prolonged immune dysregulation driven by residual bacterial products (7, 8). The present data most strongly support the residual-antigen model and refine it.

We previously showed that SARS-CoV-2 S1 protein persists in CD16⁺ monocytes in long COVID, where it is associated with chronic inflammation in the absence of ongoing viral replication (11). Similarly, FlaB and OspC persisted in CD14⁺CD16⁺ monocytes without evidence of bacterial transcriptional activity. This parallel suggests that long-lived monocyte subsets may serve as reservoirs for persistent microbial antigen across distinct post-infectious syndromes.

Three observations argue against ongoing active infection in the analyzed monocyte compartment. First, FlaB RT-qPCR was uniformly negative. Second, the bacterial reads detected were confined to structural RNA loci, with no protein-coding gene and no difference between antigen-positive and antigen-negative groups. This pattern is consistent with cross-mapping of conserved structural RNA rather than with bacterial transcription, and argues against metabolically active organisms in these samples. Third, the detected proteins were FlaB and OspC, relatively stable structural antigens rather than labile metabolic enzymes. These findings are more consistent with persistence of bacterial remnants than with ongoing replication. These data support a model in which monocytes have internalized spirochete-derived material that resists complete phagolysosomal degradation.

PBMCs from antigen-positive donors also exhibited a distinct inflammatory transcriptional program. The most strongly induced signal was TNFSF14/LIGHT, a potent mediator of inflammatory and lymphocyte-activating signaling. Antigen-positive samples further showed increased expression of GSDMD and GBP5, consistent with inflammasome engagement, along with induction of IFIT2 and IFIT3, indicating activation of an interferon-responsive program. Additional changes in CXCR1, FPR2, FCGR2B, and LILRB5 suggest broader remodeling of chemotactic and immune receptor pathways. The induction of FPR2, a receptor for bacterial N-formyl peptides, is of interest, as it offers a direct route by which retained bacterial protein could sustain myeloid activation (15). These data define an antigen-associated inflammatory state marked by inflammatory activation, interferon signaling, and inflammasome-related pathways. The greater dispersion of antigen-positive donors in principal component space is consistent with variable antigen burden between individuals (Figure 5C).

At the protein level, plasma from these participants showed inflammatory and vascular mediators above the healthy donor reference months to years after treatment, most consistently IFN-γ, IL-4 and VEGF. IL-6 is among the cytokines induced in human monocytes by live *Borrelia burgdorferi*, a response that also includes type I interferon (10). The elevation of IL-6 and IFN-γ here is concordant with the interferon- and inflammasome-associated transcriptional program observed in antigen-positive donors. The elevation of VEGF is of interest given that NCM patrol and interact with the endothelium (12). Antigen-positive participants showed higher median concentrations of IL-6, IFN-γ, TNF-α and VEGF than antigen-negative participants, in the same direction as the transcriptional and Immunogram findings, but with only three antigen-positive participants these differences cannot be statistically resolved and should be regarded as hypothesis-generating. Plasma reports the systemic milieu, not the monocyte compartment, and the participant with the highest IL-4 and VEGF concentrations was antigen-negative under the prespecified criterion, carrying FlaB at 31% coverage without OspC. Circulating cytokines and monocyte antigen status are therefore not interchangeable markers, and antigen status tracked which parts of the immune system were altered rather than how inflamed a participant was overall. This is consistent with the Immunogram result that both patient groups sat below the healthy reference overall while myeloid and inflammatory nodes rose with antigen status, indicating a redistribution of immune activity toward inflammation rather than uniformly greater immune activation.

TNFSF14/LIGHT signals through lymphotoxin beta receptor and herpesvirus entry mediator to regulate inflammatory signaling and T-cell co-stimulation, and has been implicated in chronic inflammatory disease and tissue injury (16). Its induction suggests that antigen-bearing monocytes may reinforce inflammatory crosstalk with other immune compartments.

The concurrent upregulation of GSDMD and GBP5 further points to inflammasome activation. GSDMD is the executor of pyroptotic pore formation, whereas GBP5 promotes inflammasome assembly, together supporting a state of sustained innate immune activation. Prior in vitro work has shown that heat-inactivated *Borrelia burgdorferi* is sufficient to induce TNF-α, IL-6, and IL-1β in human monocytes, providing a mechanistic precedent for inflammatory activation in the absence of viable organisms (10).

FPR2 is a G-protein-coupled receptor for N-formylated peptides, a modification largely restricted to bacterial and mitochondrial proteins, and its engagement drives chemotaxis and innate activation in myeloid cells (17). Its upregulation in antigen-positive donors offers a route by which retained bacterial protein could be sensed and could sustain myeloid activation without ongoing replication.

KCNJ2 was the most statistically significant gene and may reflect ion channel remodeling linked to persistent monocyte activation. Although its functional role in this setting remains to be established, its expression raises the possibility that antigen persistence is associated not only with inflammatory transcriptional changes but also with altered membrane signaling states that help sustain activation (18).

Genes elevated in antigen-positive donors were strongly enriched for neutrophil-associated transcripts, and this enrichment was carried by receptor and signaling genes rather than by the granule proteins that would accompany granulocyte carryover. The distinction matters: a shift in cell composition would raise the most granulocyte-restricted transcripts, such as CD177 and S100A8, which were unchanged. The pattern instead indicates remodeling of myeloid receptor programs, consistent with the elevated neutrophil and low-density neutrophil scores observed by Immunogram and arrived at through complementary compartment-level analysis.

By collapsing bulk RNA-seq into compartment-level activity scores through a curated causal network, Immunogram complements gene-level differential expression with a cell-type-oriented view of immune states that is broadly concordant with the transcriptomic separation between antigen-positive and antigen-negative donors. The most informative pattern was not a uniform increase in monocyte-associated signal in antigen-positive samples, but a divergence within the monocyte compartment: antigen-positive donors showed higher general monocyte and chronically activated macrophage scores together with reduced monocyte-activated dendritic, plasmacytoid dendritic, and myeloid dendritic/progenitor scores. The elevated monocyte and chronically activated macrophage scores parallel the concurrent upregulation of inflammasome-associated transcripts (GSDMD, GBP5) and the induction of KCNJ2 in antigen-positive samples. Collectively, these complementary observations are consistent with a state of inflammatory monocyte activation without a corresponding expansion of dendritic-cell-associated antigen-presenting programs.

The increase in CD8 terminal memory scores and the elevated activated gamma-delta subnode in antigen-positive samples extends this interpretation to the adaptive compartment. The concurrent induction of TNFSF14/LIGHT — a co-stimulator of T-cell activation through HVEM and a driver of lymphoid inflammation — provides a plausible mediator linking the inflammatory monocyte program to the terminally differentiated and innate-like adaptive features observed by Immunogram. This combined picture of innate activation, reduced antigen-presentation programs, and chronic adaptive stimulation is compatible with antigen persistence rather than active infection, because *Borrelia burgdorferi* proteins were detected in the absence of corresponding bacterial mRNA.

Notably, this chronic activation appears to come at a measurable cost to immune renewal capacity. The reduced progenitor scores in antigen-positive samples, together with reduced naive T cell, naive CD8 T cell, regulatory CD4 T cell, and naive/switchable B-cell scores, suggest that maintenance of the chronic monocyte-skewed response in PTLDS is sustained at the expense of the naive and progenitor compartments that would normally enable fresh responses to new antigenic challenge. By contrast, antigen-negative donors retained both the progenitor reservoir and the naive/regulatory pools, indicating that this loss of renewal capacity is associated specifically with monocyte-bound antigen persistence rather than being a uniform feature of PTLDS. Comparison with the external healthy reference placed this in context: both PTLDS groups showed lower activity than healthy donors across most nodes, so antigen status marked which compartments were affected rather than how severely.

The present analyses were performed on a cross-sectional cohort recruited from a single referral center. Transcriptomic, differential expression, and Immunogram analyses were performed on 6 patients, 3 per group, which limits statistical power and the precision of effect estimates. These findings are hypothesis-generating and require replication in a larger cohort. Bulk PBMC RNA-seq does not resolve which cell type contributes a given transcript, so gene-level changes cannot be attributed to monocytes directly, and the monocyte-level interpretation rests on the Immunogram deconvolution. Proteomics confirms the presence of FlaB and OspC but does not resolve their subcellular localization or degradation kinetics within monocytes. The antigen-stratified plasma comparison was limited to three antigen-positive and seven antigen-negative participants, a configuration in which no two-sided P value below 0.017 is attainable, so the directional differences observed are consistent with, but cannot independently establish, an association between monocyte antigen persistence and systemic cytokine elevation. Antigen-negative patients also experienced persistent symptoms, indicating that additional mechanisms contribute to PTLDS in some individuals.

The findings parallel previous observations in long COVID. In both syndromes, persistent microbial antigen appears to localize to CD16⁺ monocyte subsets. In long COVID, this has been linked most directly to NCM, the endothelial-patrolling monocyte subset. In the present PTLDS cohort, *Borrelia burgdorferi* FlaB and OspC were detected in sorted CD45⁺CD14⁺CD16⁺ monocytes. The transcriptomic overlap (inflammasome activation, ISG induction, TNF signaling) suggests a shared mechanism agnostic to the original pathogen. Together, these observations support a model in which post-infectious syndromes such as PTLDS and long COVID are sustained by an immune system locked in antigen-fueled dysregulation rather than by ongoing infection. Future studies will expand this work through larger, multi-center cohorts that include antigen-negative patients who remain asymptomatic. Longitudinal sampling combined with functional flow-based assays, deeper compartment-resolved immune profiling, and targeted therapeutic interventions will further define the contribution of persistent *Borrelia burgdorferi* antigens to PTLDS and accelerate the development of mechanism-based treatments for the broader class of post-infectious chronic illnesses. Given the failure of a large, randomized, controlled clinical trial looking at antibiotic intensification in PTLDS (19), therapies directed at macrophage-driven immune dysregulation may hold promise (20).

## Materials and Methods

### Patients

This study was approved by the IRB for the Chronic COVID Treatment Center (protocol CCTC-008). All participants provided written informed consent prior to participation. Following informed consent, whole blood was collected in a 10 mL EDTA tube and a 10 mL plasma preparation tube (PPT). Ten patients were identified as PTLDS based on the CDC definitions and prior publications (5, 9, 19). Patients were distinguished from long COVID using machine learning algorithms previously published (21). Donors with previous antibiotic treatment failures were recorded (Table 1). In short, inclusion criteria were:

- Prior diagnosis of Lyme disease using two-tiered or modified two-tiered testing
- Characteristic symptoms lasting longer than 6 months
- Failure of at least one cycle of appropriate antibiotics without resolution of symptoms
- Symptoms impact daily life

Exclusion criteria were a positive long hauler index (LHI) or diagnosis of ME-CFS.

Both male and female participants were included. Of the 10 patients analyzed by mass spectrometry, 4 were female and 6 were male; the RNA-sequencing subset comprised 1 female and 2 males in each group. The study was not powered to detect sex-specific effects, and sex was not included as a covariate in differential expression analysis. Findings are expected to be relevant to both sexes.

### Multiplex cytokine/chemokine profiling

Cytokine and chemokine concentrations were measured in plasma drawn into plasma preparation tubes (PPT, BD Biosciences, San Jose, CA). Each well of a 96-well plate received 25 μL of plasma, assayed with the IncellKINE 14-plex bead-based panel (InCellDx, HealthBio, Inc.) as applied previously (22). The panel quantifies IL-2, IL-4, IL-6, IL-8, IL-10, IL-13, IFN-γ, TNF-α, GM-CSF, VEGF, sCD40L, CCL3 (MIP-1α), CCL4 (MIP-1β) and CCL5 (RANTES). Beads were measured on a three-laser Beckman Coulter CytoFlex LX and analyzed in Kaluza (Beckman Coulter, Miami, FL). Concentrations are reported in pg/mL. Five analytes (IL-4, IL-6, IFN-γ, TNF-α and VEGF) were reported in this analysis. Plasma from 50 healthy donors was assayed on the same platform and panel as a reference distribution.

### Flow cytometric cell sorting

Cryopreserved PBMCs were quick-thawed, centrifuged, and washed in 2% BSA solution in D-PBS. Cells were blocked for 5 minutes in 2% BSA and then incubated at room temperature for 30 minutes with Alexa Fluor® 488 Anti-CD45 antibody (IncellDx, 1/100 dilution), 2.5 μg of Alexa Fluor® 647 Anti-CD16 antibody (BD, Cat. # 55710), and 1 μg of PerCP/Cy5.5 Anti-human CD14 antibody (Biolegend, Cat. #325622). Cells were washed twice with 2% BSA/D-PBS, filtered, and kept on ice for the duration of the cell sort. Data was acquired on a Sony SH800, and only CD45^+^ cells staining positive for both CD14^+^ and CD16^+^ were sorted into test tubes with 100 μL 2% BSA solution (Figure 2). Human blood monocytes comprise classical (CD14⁺⁺CD16⁻), IM (CD14⁺⁺CD16⁺) and NCM (CD14⁺CD16⁺⁺) subsets. This gate captures CD16⁺ monocytes and may include both IM and NCM depending on CD14 and CD16 expression intensity. Sort purity was confirmed to be >99% by reanalysis of the sorted CD45^+^CD14^+^CD16^+^ population using the same gating strategy.

### Proteomics

Sorted monocyte samples suspended in sort collection fluid were concentrated using 10 kDa Microcon centrifugal filters (Millipore, Cat. no. MRCPRT010), washed twice on-filter with water, and lysed on-filter in 5% sodium dodecyl sulfate (SDS) and NuPAGE LDS sample buffer (Thermo Fisher Scientific, Cat. no. NP0007). Lysates were reduced with NuPAGE Sample Reducing Agent (Thermo Fisher Scientific, Cat. no. NP0004) at 60 °C for 10 minutes, separated by SDS-PAGE on 4–12% Bis-Tris Mini Protein Gels (Thermo Fisher Scientific, Cat. no. NP0322BOX), and subjected to in-gel tryptic digestion using the Thermo Scientific In-Gel Tryptic Digestion Kit (Cat. no. 89871). Peptides were extracted from gel pieces with 50% acetonitrile and dried in a vacuum concentrator.

Dried peptides were reconstituted in 20 μL of 2% acetonitrile and 0.1% formic acid, and 5 μL of each sample was loaded onto EvoTip Pure tips (Evosep, EV2011) according to the manufacturer’s instructions for desalting before liquid chromatography-mass spectrometry (LC-MS) analysis. Peptides were separated on a 15 cm × 150 μm Evosep Performance column (EV1137; ReproSil-Pur C18, 1.5 μm beads) using the 30 samples per day method (44-minute gradient) and analyzed on a Thermo Orbitrap Astral mass spectrometer. Full MS scans were acquired over m/z 380–980, followed by data-independent acquisition (DIA) MS/MS using 300 non-overlapping 2 Th isolation windows spanning m/z 380.4–980.7. MS/MS spectra were acquired in the Astral analyzer over m/z 150–2000 with higher-energy collisional dissociation at 25% normalized collision energy.

### Transcriptomics

Total RNA was extracted from PBMC samples from six donors, including three antigen-negative donors (N1-N3) and three antigen-positive donors (P1-P3), using the miRNeasy Mini Kit (Qiagen) according to the manufacturer’s instructions. RNA concentration was measured using the Quant-iT RiboGreen RNA Assay (Thermo Fisher Scientific), and RNA integrity was assessed using a Fragment Analyzer (Agilent). For library preparation, 30 ng of total RNA per sample was processed using the SMARTer Stranded Total RNA-Seq Kit v2 (Takara Bio) with unique dual-index barcodes according to the manufacturer’s protocol. Library concentration was measured using the Quant-iT PicoGreen dsDNA Assay (Thermo Fisher Scientific), and library size distribution was assessed using the TapeStation system (Agilent). Libraries were further quantified by qPCR using the KAPA Library Quantification Kit (Roche). Paired-end sequencing (2 × 100 bp; approximately 100 million reads per sample) was performed on an Illumina NovaSeq X Plus instrument according to the manufacturer’s instructions.

### Borrelia flagellar filament protein (FlaB) RT-qPCR

Total RNA was quantified using a TapeStation 4200 system with the RNA ScreenTape Kit (Agilent Technologies, Santa Clara, CA, USA). Complementary DNA (cDNA) was synthesized from 500 ng of total RNA in a final volume of 12 µL using SuperScript IV Reverse Transcriptase (Thermo Fisher Scientific, Waltham, MA, USA) and random hexamer primers according to the manufacturer’s instructions. Quantitative PCR (qPCR) was then performed using 2 µL of cDNA as input. *Borrelia burgdorferi* FlaB was assayed as the target, and human RNase P was used as an internal control. The FlaB primer and probe sequences were as follows: forward, AGCTGAAGAGCTTGGAATGC; reverse, GAGTTCATGTTGGAGCAAACC; and probe, CACCAGCATCACTTTCAGGGTCTCAA. The RNase P primer and probe sequences were as follows: forward, AGATTTGGACCTGCGAGCG; reverse, GAGCGGCTGTCTCCACAAGT; and probe, TTCTGACCTGAAGGCTCTGCGCG. Amplification was performed with an initial denaturation at 95 °C for 10 minutes, followed by 40 cycles of 95 °C for 15 seconds and 60 °C for 30 seconds.

### Data Analysis

#### Proteomics

Data were uploaded to the Proteome Discoverer 3.1 (Thermo Scientific) and searched in Chimerys 2.0 search algorithm using combined human and Borrelia FASTA database that included reviewed canonical and isoform entries (downloaded August 26, 2024). Fixed modifications included carbamidomethylation on cysteine. Processing mode was ‘One job for All files’ and Prediction model was ‘Inferys_2.1.1_fragmentation’. Validation was set to target FDR strict: 0.01, target FDR relaxed: 0.05.

#### Quality control analysis

To evaluate LC-MS performance, technical replicates of Pierce™ HeLa Protein Digest Standard (Thermo Fisher Scientific, Cat Num: 88329) were analyzed regularly prior, post and between sample queues. 200 ng of HeLa protein digests were separated on the LC column and analyzed using the methods described above. Acquired files were searched in PAS using the DIANN (1.8.1) search engine against the system-provided human spectral library. Precursor and protein group false discovery rates were both controlled at 1%. Over 10,000 protein groups were identified across all HeLa samples, and the coefficient of variation of the number identified was less than 0.1%. In addition, samples containing 80 fmol (on column) of PepCalMix (Sciex, P/N: 5045757) were analyzed for every three samples. PepCalMix data were analyzed in Skyline. Retention time and Peptide Peak Area of the PepCalMix were used to check the LC and MS performance throughout the sample acquisition. These results confirmed the performance of the LC-MS throughout the project.

#### Transcriptomics

Paired-end reads were preprocessed with fastp v1.3.7 using automatic adapter detection for paired-end reads, poly-G and poly-X tail trimming, 3′ end quality trimming, a Phred quality threshold of 20, removal of reads with more than 40% unqualified bases, and a minimum read length of 36 bp after trimming. Trimmed reads were pseudoaligned to the Homo sapiens GRCh38 cDNA reference (Ensembl release 115; 328,868 transcripts) using kallisto v0.51.1 with reverse-strand orientation. Transcript-level estimates were aggregated to gene-level counts via Ensembl BioMart mappings.

Differential expression of PBMC genes was performed with PyDESeq2 (negative-binomial Wald test, median-of-ratios normalization) across all 19,744 testable genes, with Benjamini–Hochberg adjustment across the full set. Figures and gene rankings were restricted to genes with evidence of expression in normal blood, assessed using an external healthy reference of sorted immune-cell populations (GSE107011) (23). A gene was retained if it reached ≥5 transcripts per million (TPM) in at least one reference subset. This retained 14,741 of 19,744 genes. Because the restriction was applied after multiple-testing correction, reported P_adj_ values remain conservative.

Cell-type enrichment among differentially expressed genes was assessed by assigning each gene to the sorted immune-cell population with the highest median expression and comparing the resulting distribution against blood-expressed genes using Fisher’s exact test.

To assess bacterial transcription, reads were separately quantified against the *Borrelia burgdorferi* B31 reference genome and annotation (GCF_000008685.2, ASM868v2) using Salmon, with reads mapped to the bacterial reference independently of the human alignment. Differential expression between antigen-positive and antigen-negative samples was performed with DESeq2, with Benjamini–Hochberg adjustment applied across the ten detected bacterial loci (Figure S2).

#### Immunogram analysis

Bulk RNA-seq data from PBMCs were aligned and converted to raw gene counts. Raw counts were analyzed with the Immunogram pipeline (Caucell Inc) (24, 25). Counts were normalized, batch-corrected, and log-transformed. Features were then projected onto a pre-trained probabilistic graphical model (directed acyclic graph) constructed from >10,000 curated causal interactions among immune genes, signaling pathways, and cell states. Bayesian network inference was used to propagate the observed transcriptional perturbations through the full causal network, yielding for each node a posterior activity score that estimates how strongly the corresponding immune gene, pathway, or cell state is engaged in the sample.

Node-level activity scores were organized into one system-level Immunogram, which collapsed high-dimensional node activities into 16 cross-compartment immune axes, and seven compartment-specific sub-Immunograms representing monocyte, granulocyte, natural-killer, T-invariant/gamma-delta T-cell, CD4 T-cell, CD8 T-cell, and B-cell lineages. Immunogram profiles were compared between antigen-positive and antigen-negative PTLDS participants. Thirteen external healthy PBMC RNA-seq samples from GSE107011 were additionally processed through the same workflow and displayed as a descriptive reference cohort. Because these healthy samples originated from an independent study, they were not included in differential-expression analyses or formal statistical comparisons.

Posterior activity scores were generated on a 0-1 scale within the Immunogram framework. Given three participants per group, Immunogram comparisons are descriptive and reported as consistent directional shifts in node-level activity rather than as statistically tested effects. For visualization of the three-group analyses, radar and bubble plots display activity scores to contextualize antigen-positive and antigen-negative profiles against the external healthy reference. Each Immunogram was rendered as a radar plot accompanied by a bubble plot of the corresponding Immunogram activity scores.

### Statistics

All tests were two-sided, with uncorrected P < 0.05 considered nominally significant. Plasma analyte concentrations were compared against the healthy donor median by exact sign test, and between antigen-positive and antigen-negative participants by exact Mann–Whitney U test. Neither test assumes normality or is affected by individual extreme values. With three antigen-positive and seven antigen-negative participants the smallest attainable two-sided Mann–Whitney P value is 0.017, so those comparisons are reported as effect directions rather than hypothesis tests. Analyte P values were Bonferroni-corrected across the five analytes, with Padj < 0.05 considered significant, corresponding to an uncorrected P < 0.010. Differential gene expression was assessed with PyDESeq2 using a negative-binomial Wald test and Benjamini–Hochberg adjustment across all 19,744 testable genes, then restricted to genes reaching ≥5 TPM in at least one sample of a healthy immune-cell reference (GSE107011), a criterion set before examining these results. Of the 14,741 genes retained, 213 were differentially expressed at Padj < 0.05, compared with 307 before the restriction. Enrichment of neutrophil-characteristic genes was tested by two-sided Fisher’s exact test against the remainder of that set, in which 8.6% were neutrophil-characteristic. Immunogram comparisons were descriptive and not statistically tested, given three participants per group.

## Data availability

The data supporting the findings of this study are available within this article and in the supplemental material. The RNA sequencing data discussed have been deposited in NCBI’s Gene Expression Omnibus (Patterson *et al*., 2026) and are accessible through GEO Series accession number GSE348165.

## Acknowledgments

This research received no specific grant from any funding agency in the public, commercial, or not-for-profit sectors. Kwat Medetgul-Ernar was supported by the Stanford University Medical Scientist Training Program grant T32-GM007365. BKP conceptualized and designed the experiment and supervised the study. RY recruited and enrolled patients. EBF performed cell and protein analysis experiments. CB, KB, PAF analyzed data. NC analyzed data and prepared the manuscript. KM-E analyzed data and prepared Immunogram results. All authors reviewed and approved the manuscript.

---

**Figure S1.**
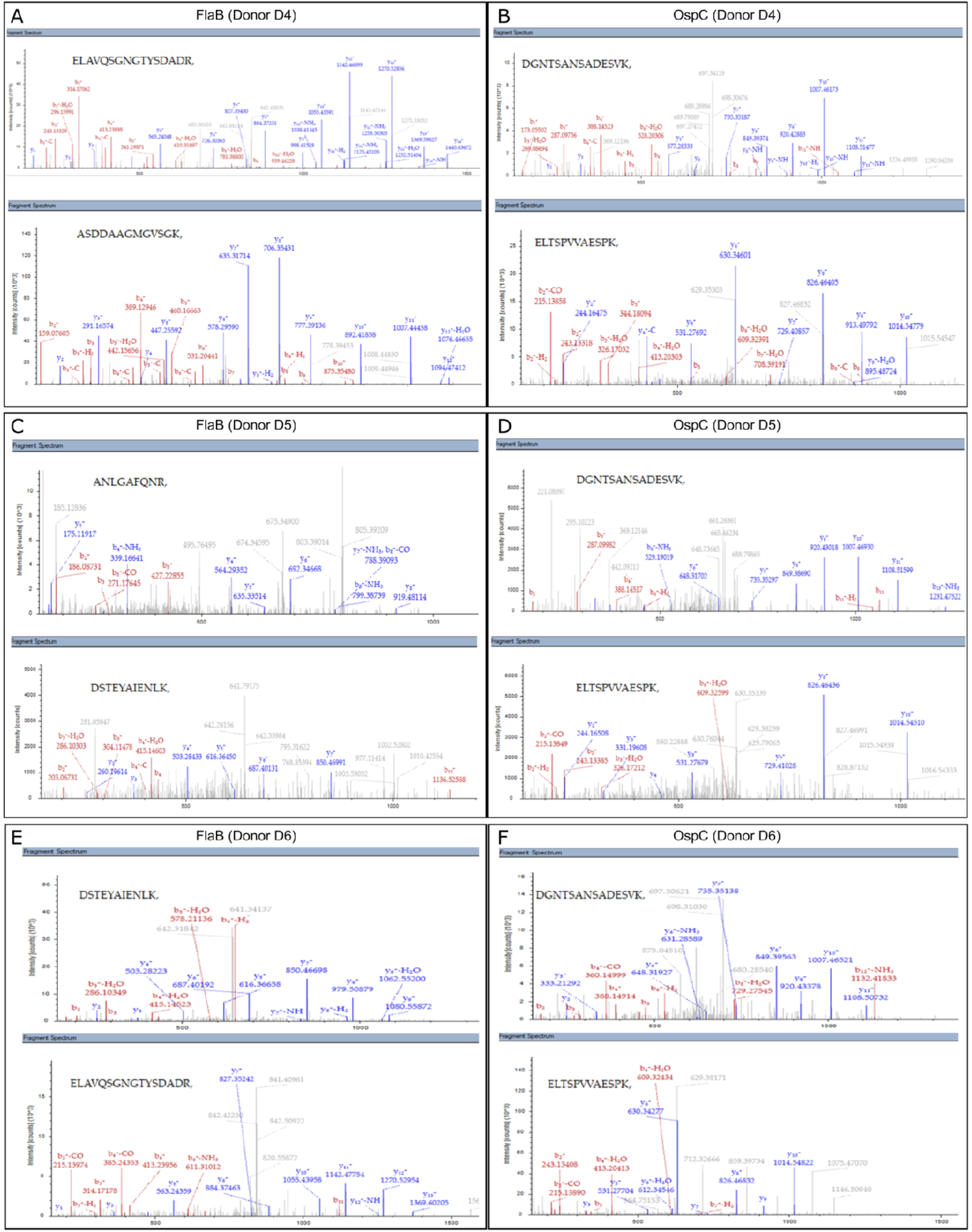
Representative LC–MS/MS fragment ion spectra of selected peptides identified in samples D4 (A, B), D5 (C, D), and D6 (E, F) from FlaB (A, C, E) and OspC (B, D, F).

**Figure S2.**
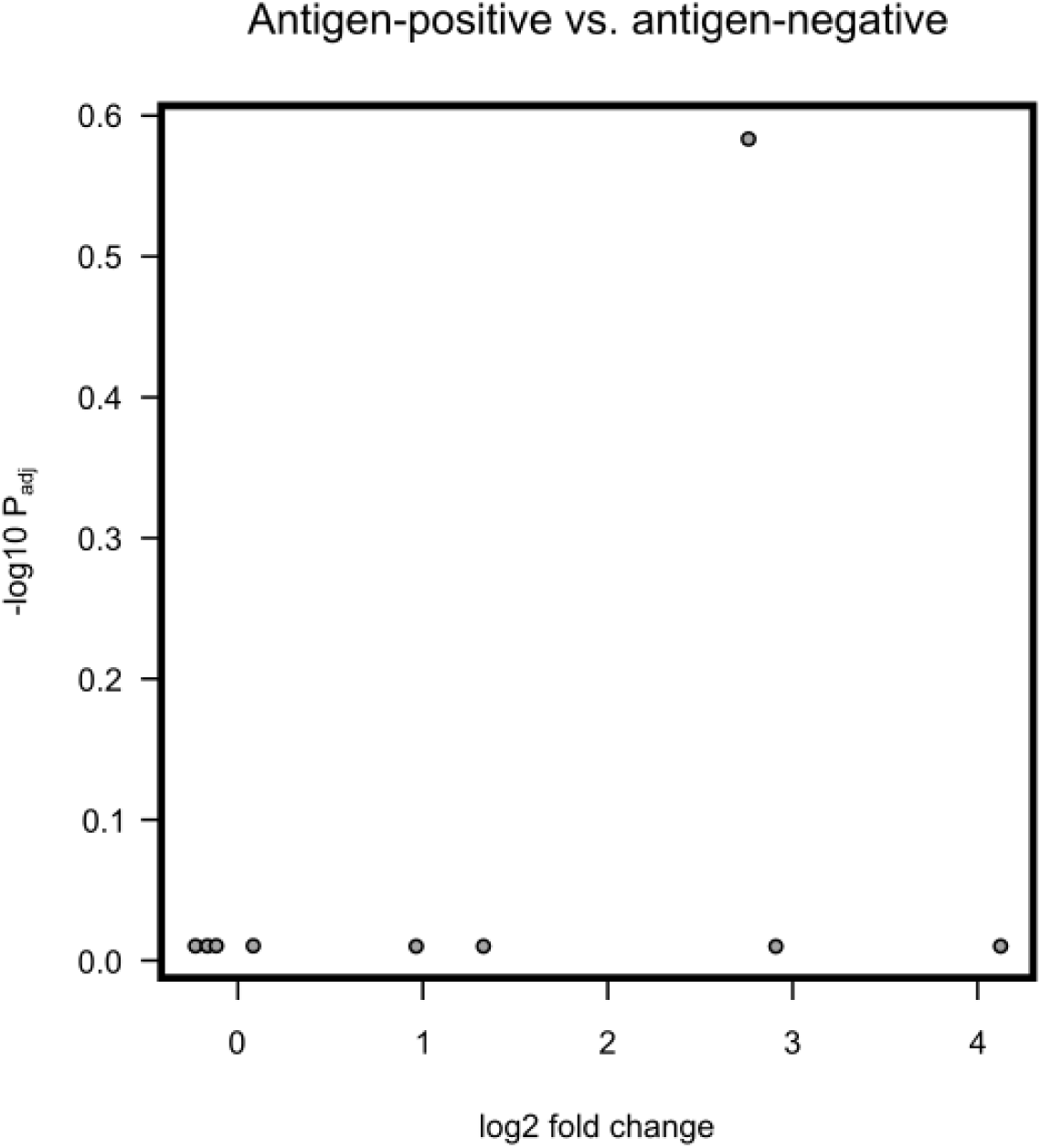
Differential expression of *Borrelia burgdorferi* loci detected in PBMC RNA-seq. Reads were quantified against the *Borrelia burgdorferi* B31 reference genome and tested for differential abundance between antigen-positive and antigen-negative samples, with positive log₂ fold change indicating higher abundance in antigen-positive samples. All ten detected loci were non-coding structural RNAs — ribosomal RNA, transfer RNA, and tmRNA — and no protein-coding bacterial gene was detected. Adjusted P values were computed across these ten loci and none reached significance, the lowest being P_adj_ = 0.26.

**Figure S3.**
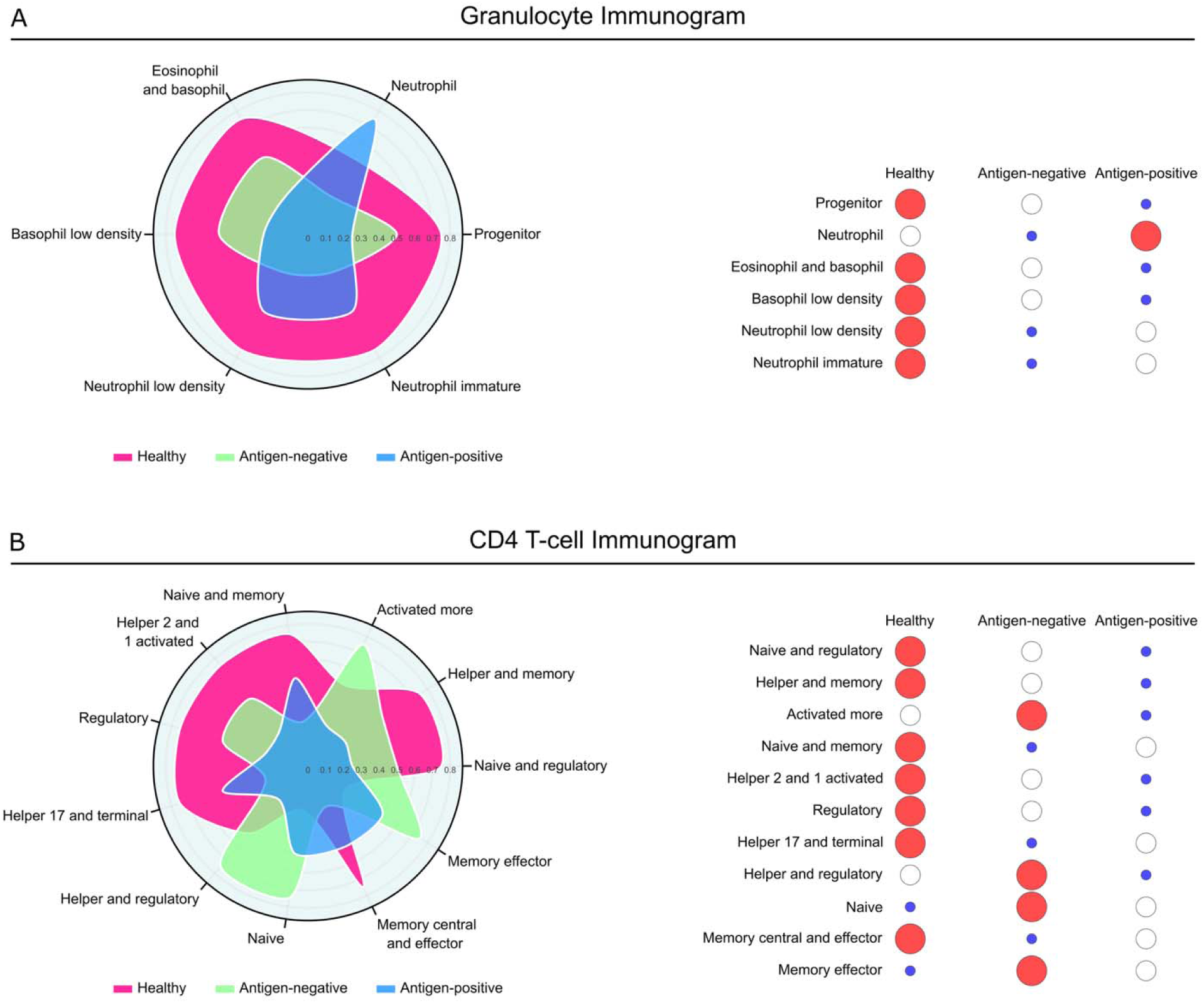
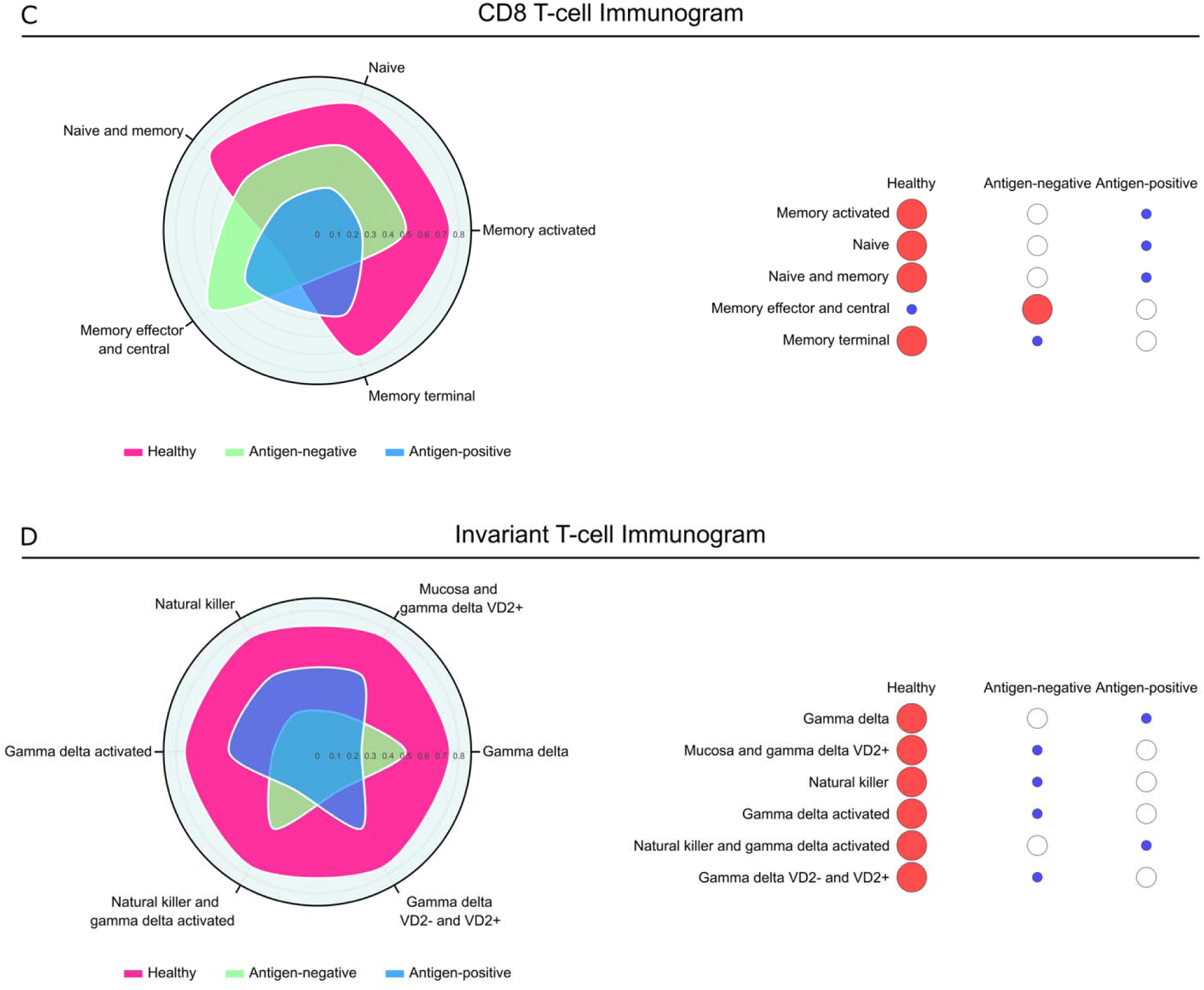
Additional sub-Immunograms. In antigen-positive samples: (A) granulocyte — elevated neutrophil and low-density neutrophil scores; (B) CD4 T cell — reduced naive and regulatory CD4 T-cell scores; (C) CD8 T cell — elevated terminal memory scores together with reduced naive CD8 T-cell scores; (D) T invariant / gamma-delta — elevated activated gamma-delta subnode; (E) B cell — elevated plasmablast scores together with reduced plasma cell, progenitor, and naive/switchable B-cell scores; (F) natural killer — elevated activated natural killer scores. Each panel shows a radar plot (left) and a bubble plot (right) of the same per-axis activity scores on a 0–1 scale. Antigen-negative samples are shown in green, antigen-positive samples in blue, and an external healthy reference cohort (GSE107011, N = 13) in magenta. The healthy cohort is shown for context only and was not included in statistical comparisons.

**Table S1.** Mass spectrometry performance of negative control and protein standards. Sequence coverage, peptide-spectrum matches, and peptide counts for the negative control, FlaB standard, and OspC standard analyzed alongside study samples. Both standards were prepared from whole *Borrelia burgdorferi* lysate and therefore contain both antigens. The absence of either protein in the negative control confirms that detection in patient samples did not arise from carryover (not detected, ND).

| Control | FlaB |  |  | OspC |  |  |
| --- | --- | --- | --- | --- | --- | --- |
|  | Coverage (%) | Number of peptide spectrum matches | Number of peptides | Coverage (%) | Number of peptide spectrum matches | Peptides |
| Negative control | ND | ND | ND | ND | ND | ND |
| FlaB standard | 60% | 7079 | 32 | 35% | 33 | 8 |
| OspC standard | 48% | 1269 | 13 | 86% | 4929 | 32 |

**Table S2.** RT-qPCR results for *Borrelia burgdorferi* FlaB and human RNase P in RNA-seq samples. All six RNA-seq samples were negative for FlaB by RT-qPCR. RNase P cycle threshold values are shown as an internal control for amplifiable host nucleic acid. Donor labels N1–N3 and P1–P3 denote antigen-negative and antigen-positive RNA-seq samples, respectively.

| Donor ID | FlaB qPCR result | Human RP qPCR (cycle threshold) |
| --- | --- | --- |
| D1 (N1) | Negative | 26.53 |
| D2 (N2) | Negative | 27.00 |
| D3 (N3) | Negative | 26.41 |
| D4 (P1) | Negative | 26.15 |
| D5 (P2) | Negative | 25.67 |
| D6 (P3) | Negative | 26.04 |

**Table S3.** Differentially expressed genes between antigen-positive and antigen-negative donors (P_adj_ < 0.05). Genes are grouped by direction — those higher in antigen-positive (P) donors first, then those higher in antigen-negative (N) donors — and ordered by adjusted P within each group. Positive log₂ fold change indicates higher expression in antigen-positive donors. Mean expression is the average normalized count across all six samples. Maximum TPM in healthy blood is each gene’s highest value across the samples of the healthy immune-cell reference (GSE107011). The most characteristic cell type is the reference subset in which the gene’s median expression across replicate samples is highest. Assignment indicates where a transcript is most abundant in healthy blood and does not imply lineage exclusivity, since many assigned genes are expressed by several populations. Reference subsets are neutrophils, basophils, classical, intermediate and non-classical monocytes (C_mono, I_mono, NC_mono), myeloid and plasmacytoid dendritic cells (mDC, pDC), natural killer cells (NK), hematopoietic progenitors (Progenitor), naive, non-switched memory, switched memory and exhausted B cells (B_naive, B_NSM, B_SM, B_Ex), plasmablasts, naive and terminal effector CD4 T cells (CD4_naive, CD4_TE), the T helper subsets Th1, Th2, Th17 and Th1/Th17, follicular helper (TFH) and regulatory (Treg) CD4 T cells, naive, central memory, effector memory and terminal effector CD8 T cells (CD8_naive, CD8_CM, CD8_EM, CD8_TE), mucosal-associated invariant T cells (MAIT), and Vδ2⁺ and Vδ2^-^ γδ T cells (VD2+, VD2−).

| Gene | Mean expression | log2 fold change | Standard error of log2 fold change | P | Padj | Higher in | Maximum TPM in healthy blood | Most characteristic cell type |
| --- | --- | --- | --- | --- | --- | --- | --- | --- |
| KCNJ2 | 655.3 | 3.317 | 0.354 | 7.78E-21 | 1.71E-17 | P | 75.9 | Neutrophils |
| TNFSF14 | 1173 | 1.969 | 0.245 | 8.92E-16 | 1.36E-12 | P | 72.7 | Neutrophils |
| CCDC121 | 107.5 | 2.644 | 0.362 | 2.86E-13 | 3.32E-10 | P | 7.7 | Basophils |
| FMC1-LUC7L2 | 60 | 6.193 | 0.861 | 6.57E-13 | 7.2E-10 | P | 5.3 | CD4_TE |
| DSP | 1110.1 | 2.704 | 0.436 | 5.56E-10 | 0.000000477 | P | 19.5 | B_naive |
| B3GALT4 | 736.3 | 1.314 | 0.238 | 3.41E-08 | 0.000025 | P | 56.1 | NK |
| ORMDL3 | 2033.6 | 0.999 | 0.182 | 4.27E-08 | 0.0000301 | P | 212.7 | CD8_EM |
| SIGLEC5 | 2786.4 | 1.47 | 0.273 | 7.38E-08 | 0.0000486 | P | 171.9 | Neutrophils |
| BBS12 | 117.3 | 1.423 | 0.269 | 0.000000121 | 0.0000723 | P | 6.5 | Progenitor |
| KRT23 | 534.7 | 3.856 | 0.826 | 0.00000303 | 0.00115 | P | 317.4 | Neutrophils |
| FADD | 743.9 | 1.485 | 0.32 | 0.00000341 | 0.00125 | P | 32.1 | Neutrophils |
| PLEKHF1 | 427.9 | 1.317 | 0.285 | 0.00000383 | 0.00138 | P | 165.6 | NK |
| MGAM2 | 163.7 | 4.975 | 1.079 | 0.000004 | 0.00139 | P | 15.7 | Neutrophils |
| NCF1B | 3962.2 | 1.824 | 0.395 | 0.00000394 | 0.00139 | P | 915.3 | Neutrophils |
| GSDMD | 3066.5 | 0.795 | 0.173 | 0.00000447 | 0.00151 | P | 191.8 | CD8_EM |
| RGS3 | 3197.9 | 1.025 | 0.224 | 0.00000453 | 0.00151 | P | 353.1 | NK |
| TMEM234 | 1030.4 | 0.796 | 0.174 | 0.0000046 | 0.00151 | P | 41.6 | Progenitor |
| CDC42EP2 | 1125.2 | 1.311 | 0.29 | 0.00000605 | 0.00193 | P | 122.2 | Neutrophils |
| CXCR1 | 731 | 3.511 | 0.787 | 0.00000813 | 0.00236 | P | 2144.3 | Neutrophils |
| TRADD | 1265.3 | 0.79 | 0.177 | 0.00000813 | 0.00236 | P | 180.8 | TFH |
| TAF1D | 22114 | 0.825 | 0.185 | 0.00000838 | 0.0024 | P | 386.2 | Progenitor |
| CLEC18A | 239.8 | 2.177 | 0.497 | 0.0000116 | 0.00314 | P | 6 | Neutrophils |
| TMEM51 | 171.7 | 1.616 | 0.369 | 0.0000118 | 0.00315 | P | 5.1 | C_mono |
| ARHGEF5 | 474.5 | 0.815 | 0.188 | 0.0000145 | 0.00358 | P | 7.1 | B_NSM |
| LAIR2 | 108.5 | 2.372 | 0.549 | 0.0000155 | 0.00372 | P | 52 | NK |
| FCGR2B | 3880.9 | 1.234 | 0.286 | 0.0000161 | 0.00383 | P | 551.1 | mDC |
| CHD8 | 33662.5 | 0.606 | 0.141 | 0.000017 | 0.004 | P | 69.1 | CD8_naive |
| ZNF213 | 648.9 | 0.712 | 0.166 | 0.0000173 | 0.00402 | P | 9.3 | Progenitor |
| ACD | 668.5 | 0.691 | 0.162 | 0.000019 | 0.00437 | P | 77 | VD2- |
| FPR2 | 1138 | 1.743 | 0.408 | 0.0000193 | 0.00438 | P | 1261.1 | Neutrophils |
| TFEB | 3819 | 0.681 | 0.16 | 0.0000209 | 0.00451 | P | 145 | B_SM |
| RPP25L | 206.3 | 1.003 | 0.237 | 0.0000231 | 0.00479 | P | 30.1 | Th1 |
| KIAA0319 | 149.5 | 2.769 | 0.656 | 0.0000242 | 0.00487 | P | 5.9 | Neutrophils |
| KBTBD7 | 609.8 | 1.038 | 0.246 | 0.000025 | 0.00498 | P | 53.9 | Neutrophils |
| LILRB5 | 658.9 | 1.317 | 0.315 | 0.0000281 | 0.00555 | P | 7.6 | Neutrophils |
| EML2 | 9085.5 | 0.861 | 0.206 | 0.0000295 | 0.00577 | P | 125 | NK |
| IFT70B | 182.2 | 1.027 | 0.247 | 0.0000321 | 0.00604 | P | 10.3 | Progenitor |
| MYBPH | 81.3 | 4.433 | 1.069 | 0.0000339 | 0.00619 | P | 37.2 | pDC |
| RPL30P7 | 27.5 | 3.908 | 0.953 | 0.0000413 | 0.00716 | P | 10.8 | Neutrophils |
| SPSB2 | 186.1 | 1.562 | 0.382 | 0.0000443 | 0.00761 | P | 71.2 | Th1 |
| ZNF365 | 105 | 2.116 | 0.52 | 0.0000474 | 0.00808 | P | 19.7 | CD4_TE |
| COL9A3 | 109 | 2.3 | 0.567 | 0.0000502 | 0.00845 | P | 97.6 | B_NSM |
| S1PR1 | 3885.4 | 0.969 | 0.239 | 0.0000507 | 0.00845 | P | 759.2 | Basophils |
| YJU2B | 2750.8 | 0.632 | 0.157 | 0.0000571 | 0.00924 | P | 114.7 | CD8_TE |
| HDC | 65 | 2.302 | 0.575 | 0.0000624 | 0.00993 | P | 11144.4 | Basophils |
| TJAP1 | 2757.3 | 0.557 | 0.141 | 0.0000728 | 0.0111 | P | 34.9 | Th2 |
| BTN3A1 | 8970.6 | 0.818 | 0.207 | 0.0000789 | 0.0114 | P | 392.2 | VD2- |
| CASQ1 | 44.9 | 3.751 | 0.949 | 0.0000771 | 0.0114 | P | 8.7 | mDC |
| CISH | 1362.2 | 1.244 | 0.315 | 0.0000785 | 0.0114 | P | 584.7 | Basophils |
| RPS10-NUDT3 | 162.9 | 1.131 | 0.287 | 0.0000794 | 0.0114 | P | 29.1 | Progenitor |
| TNFRSF10C | 5001.9 | 2.843 | 0.721 | 0.000081 | 0.0114 | P | 2196.3 | Neutrophils |
| TRPM6 | 787.2 | 3.104 | 0.786 | 0.0000777 | 0.0114 | P | 74 | Basophils |
| ZNF658 | 404.7 | 1.117 | 0.283 | 0.0000813 | 0.0114 | P | 13.9 | Neutrophils |
| SH2B2 | 1210.6 | 1.022 | 0.26 | 0.0000837 | 0.0116 | P | 22.1 | Neutrophils |
| ASS1P1 | 176.5 | 1.513 | 0.387 | 0.0000925 | 0.0123 | P | 37.5 | Progenitor |
| FCRL5 | 3819.8 | 1.186 | 0.303 | 0.0000914 | 0.0123 | P | 373.6 | B_Ex |
| ISY1-RAB43 | 1189.5 | 0.615 | 0.157 | 0.0000938 | 0.0124 | P | 10.7 | CD4_naive |
| CTBP2P8 | 140 | 2.823 | 0.724 | 0.0000958 | 0.0126 | P | 13.4 | Neutrophils |
| MEIS1 | 481 | 1.118 | 0.287 | 0.0000972 | 0.0127 | P | 984.9 | Progenitor |
| LINC02218 | 81.7 | 4.24 | 1.088 | 0.0000979 | 0.0127 | P | 31.3 | Neutrophils |
| IFIT1 | 383.9 | 0.95 | 0.244 | 0.0000987 | 0.0127 | P | 1837.1 | Neutrophils |
| C1orf35 | 1776.3 | 0.557 | 0.143 | 0.0001 | 0.0129 | P | 20.2 | Th2 |
| SLC1A7 | 50.8 | 1.869 | 0.481 | 0.000101 | 0.0129 | P | 31.6 | NK |
| GBP5 | 9864.8 | 1.212 | 0.313 | 0.000108 | 0.0134 | P | 1844.8 | MAIT |
| RAB24 | 5878.6 | 0.93 | 0.241 | 0.000111 | 0.0136 | P | 443.9 | NC_mono |
| FCHO1 | 5612.5 | 0.505 | 0.131 | 0.000117 | 0.0142 | P | 114.5 | Neutrophils |
| BTNL8 | 476.7 | 3.219 | 0.837 | 0.000121 | 0.0144 | P | 170.8 | Neutrophils |
| H4C15 | 987.9 | 1.713 | 0.446 | 0.000123 | 0.0146 | P | 75.8 | Neutrophils |
| GAPDHP14 | 43.9 | 2.442 | 0.637 | 0.000125 | 0.0148 | P | 8.4 | Neutrophils |
| HSPA6 | 1499.5 | 1.828 | 0.478 | 0.000133 | 0.0155 | P | 334.4 | Neutrophils |
| KCNJ15 | 3147.2 | 2.575 | 0.674 | 0.000134 | 0.0155 | P | 890 | Neutrophils |
| SLC35E2A | 328.7 | 1.3 | 0.34 | 0.000134 | 0.0155 | P | 24.2 | Progenitor |
| CHIT1 | 161.7 | 1.481 | 0.391 | 0.000149 | 0.017 | P | 18 | Neutrophils |
| ITPKC | 969.3 | 0.94 | 0.248 | 0.00015 | 0.017 | P | 5.8 | CD4_TE |
| SH3BP5L | 2655.8 | 1.118 | 0.296 | 0.000156 | 0.0173 | P | 17.8 | Basophils |
| AGAP7P | 93.8 | 3.336 | 0.884 | 0.00016 | 0.0175 | P | 53.5 | Neutrophils |
| HDDC2 | 897.4 | 0.774 | 0.206 | 0.000169 | 0.0181 | P | 116.6 | NK |
| PFDN6 | 1572.4 | 2.08 | 0.553 | 0.000169 | 0.0181 | P | 11.9 | pDC |
| ZRSR2P1 | 89.1 | 10.15 | 2.701 | 0.000171 | 0.0182 | P | 7.4 | Th1/Th17 |
| STMN1P1 | 219.2 | 1.023 | 0.275 | 0.000196 | 0.0204 | P | 8 | B_NSM |
| SUMO1P1 | 108.3 | 4.114 | 1.105 | 0.000196 | 0.0204 | P | 92.8 | Neutrophils |
| AFAP1L2 | 149.7 | 1.008 | 0.272 | 0.000209 | 0.0214 | P | 9.3 | Progenitor |
| KRT8P34 | 314.3 | 1.361 | 0.367 | 0.000211 | 0.0214 | P | 16.5 | B_NSM |
| ZNF646 | 4278.8 | 0.841 | 0.227 | 0.000214 | 0.0216 | P | 21.4 | Basophils |
| GOLGA6L5P | 788.6 | 0.85 | 0.231 | 0.000234 | 0.0233 | P | 11.7 | Progenitor |
| RTEL1P1 | 757.7 | 1.944 | 0.53 | 0.000242 | 0.0239 | P | 8.9 | Neutrophils |
| FIS1 | 2228.8 | 0.564 | 0.154 | 0.000245 | 0.024 | P | 97.6 | VD2+ |
| NCF1 | 16874.8 | 1.185 | 0.323 | 0.000247 | 0.0241 | P | 3242.3 | Neutrophils |
| MGAM | 3659.2 | 4.125 | 1.126 | 0.00025 | 0.0243 | P | 205.4 | Neutrophils |
| CD274 | 233.1 | 1.001 | 0.276 | 0.000281 | 0.0269 | P | 126 | Basophils |
| CYB5D2 | 491.4 | 0.687 | 0.189 | 0.000285 | 0.0272 | P | 27.5 | Progenitor |
| ALG1L13P | 76.6 | 3.215 | 0.887 | 0.000289 | 0.0273 | P | 147.7 | Neutrophils |
| LIMK2 | 5985.9 | 2.067 | 0.57 | 0.000289 | 0.0273 | P | 505.4 | Neutrophils |
| EXOSC4 | 528 | 1.225 | 0.338 | 0.000291 | 0.0274 | P | 24.5 | Neutrophils |
| PSMB9 | 3595.5 | 0.819 | 0.227 | 0.000302 | 0.0281 | P | 653.1 | Neutrophils |
| DYNLRB2 | 12.1 | 4.761 | 1.331 | 0.000347 | 0.0315 | P | 5.9 | Progenitor |
| MOCS3 | 360.2 | 0.962 | 0.27 | 0.000357 | 0.0317 | P | 7.8 | Progenitor |
| TRAV12-3 | 210.5 | 1.227 | 0.344 | 0.000357 | 0.0317 | P | 573.7 | TFH |
| TMEM187 | 181.6 | 0.904 | 0.254 | 0.000361 | 0.032 | P | 31.4 | TFH |
| HDGFL2 | 2704.2 | 0.514 | 0.144 | 0.000363 | 0.032 | P | 8.6 | Progenitor |
| B3GNT8 | 317.6 | 1.991 | 0.56 | 0.000381 | 0.0327 | P | 96.7 | Neutrophils |
| TMEM60 | 145.5 | 1.077 | 0.303 | 0.000379 | 0.0327 | P | 78.5 | Basophils |
| LTB | 3066.3 | 0.611 | 0.172 | 0.000384 | 0.0328 | P | 1320.6 | B_NSM |
| DEF8 | 7271.3 | 0.658 | 0.186 | 0.000391 | 0.0331 | P | 219.9 | Neutrophils |
| NCF1C | 3487.3 | 1.458 | 0.411 | 0.000393 | 0.0331 | P | 766.2 | Neutrophils |
| SOWAHD | 275.5 | 1.696 | 0.479 | 0.000395 | 0.0331 | P | 20.7 | B_SM |
| ZER1 | 2659.6 | 0.573 | 0.162 | 0.000409 | 0.0339 | P | 88.5 | Basophils |
| KIF28P | 112.8 | 0.991 | 0.282 | 0.000432 | 0.0355 | P | 99.3 | Basophils |
| DBN1 | 1397.2 | 0.948 | 0.271 | 0.000461 | 0.0376 | P | 55.3 | Progenitor |
| TEX101 | 165.7 | 1.031 | 0.294 | 0.000462 | 0.0376 | P | 124.6 | Basophils |
| SLC4A4 | 360.1 | 0.843 | 0.241 | 0.000467 | 0.0378 | P | 6.1 | NK |
| CNTNAP3B | 60.1 | 2.405 | 0.69 | 0.00049 | 0.0378 | P | 16.5 | Neutrophils |
| IFIT2 | 2599.5 | 1.724 | 0.494 | 0.000483 | 0.0378 | P | 4393.4 | Neutrophils |
| MED18 | 305.5 | 0.803 | 0.23 | 0.000487 | 0.0378 | P | 21 | Neutrophils |
| POLD4 | 2097.3 | 0.555 | 0.159 | 0.00049 | 0.0378 | P | 316.1 | B_naive |
| PSMC3IP | 263.4 | 0.967 | 0.277 | 0.000473 | 0.0378 | P | 14.3 | CD4_TE |
| SMIM35 | 159 | 0.766 | 0.22 | 0.000501 | 0.0382 | P | 10 | Treg |
| CNTNAP3 | 1334.7 | 2.474 | 0.712 | 0.000507 | 0.0384 | P | 98.4 | Neutrophils |
| DYRK1B | 916.2 | 0.649 | 0.187 | 0.000508 | 0.0384 | P | 52.4 | Progenitor |
| LILRB3 | 7964.7 | 1.347 | 0.388 | 0.000523 | 0.0393 | P | 1402.4 | Neutrophils |
| PPP1R10 | 10918.4 | 0.812 | 0.234 | 0.000523 | 0.0393 | P | 112 | C_mono |
| THBD | 1090.8 | 1.645 | 0.474 | 0.000528 | 0.0395 | P | 62.2 | Neutrophils |
| CCNJL | 1290.7 | 1.858 | 0.537 | 0.000544 | 0.0397 | P | 46.3 | Neutrophils |
| IFIT3 | 1568.5 | 1.248 | 0.361 | 0.000544 | 0.0397 | P | 3001.9 | Neutrophils |
| IGHV3-48 | 45.7 | 3.155 | 0.912 | 0.000542 | 0.0397 | P | 6385.4 | Plasmablasts |
| PRKD2 | 7064.4 | 0.55 | 0.159 | 0.000547 | 0.0397 | P | 123.7 | Neutrophils |
| ZNF613 | 690.8 | 0.636 | 0.184 | 0.00055 | 0.0397 | P | 21.5 | Progenitor |
| LSM10 | 1280.6 | 0.598 | 0.174 | 0.000599 | 0.0423 | P | 126.3 | B_naive |
| HMGB1P5 | 1176.1 | 0.724 | 0.211 | 0.000602 | 0.0423 | P | 144 | Progenitor |
| AVIL | 614.4 | 1.585 | 0.464 | 0.000643 | 0.0443 | P | 39.2 | Neutrophils |
| ADGRE3 | 1405.3 | 1.829 | 0.536 | 0.00065 | 0.0444 | P | 472.2 | Neutrophils |
| BATF2 | 75.8 | 1.547 | 0.454 | 0.000663 | 0.0451 | P | 39 | Neutrophils |
| MPZ | 264.4 | 1.395 | 0.41 | 0.000681 | 0.0462 | P | 10 | Neutrophils |
| MRPL18 | 914.3 | 0.713 | 0.21 | 0.00069 | 0.0467 | P | 69.5 | Treg |
| CRK | 2946.4 | 0.674 | 0.199 | 0.000693 | 0.0467 | P | 37.5 | Neutrophils |
| TAF11L2 | 22.7 | 4.911 | 1.448 | 0.000696 | 0.0468 | P | 6.1 | Neutrophils |
| TREML2 | 1697.2 | 1.243 | 0.367 | 0.000701 | 0.0469 | P | 159.9 | Neutrophils |
| ARL11 | 1856.9 | 0.998 | 0.296 | 0.000736 | 0.0485 | P | 54 | Neutrophils |
| HEY1 | 18.7 | 3.599 | 1.066 | 0.000733 | 0.0485 | P | 9 | Neutrophils |
| NUDC | 4859.4 | 0.478 | 0.142 | 0.000744 | 0.0485 | P | 123.4 | pDC |
| RGS14 | 4470.1 | 0.978 | 0.29 | 0.000737 | 0.0485 | P | 212.6 | Neutrophils |
| ZNF467 | 4571.4 | 0.981 | 0.291 | 0.00075 | 0.0487 | P | 74.3 | Neutrophils |
| TATDN2 | 4057.8 | 0.424 | 0.126 | 0.000764 | 0.0495 | P | 22.9 | NK |
| HTRA2 | 539.7 | 0.754 | 0.224 | 0.000775 | 0.0499 | P | 74.6 | Basophils |
| DTNA | 1135.1 | -1.89 | 0.226 | 6.85E-17 | 1.23E-13 | N | 31.7 | mDC |
| PCSK5 | 72266.6 | -2.915 | 0.394 | 1.47E-13 | 1.93E-10 | N | 60.3 | CD4_naive |
| HLA-DQB1 | 27417.2 | -1.232 | 0.236 | 0.00000017 | 0.0000932 | N | 3949.8 | mDC |
| RBM44 | 587.5 | -0.981 | 0.198 | 0.000000698 | 0.000336 | N | 5.1 | CD4_TE |
| COL24A1 | 425.8 | -1.261 | 0.259 | 0.00000113 | 0.000496 | N | 31 | Progenitor |
| MT-ND1 | 188839.4 | -0.795 | 0.164 | 0.00000135 | 0.000567 | N | 4157.4 | PBMC |
| PAICS | 1777.8 | -0.774 | 0.16 | 0.00000139 | 0.000571 | N | 139.8 | Progenitor |
| PDE10A | 656.1 | -0.823 | 0.183 | 0.00000718 | 0.00221 | N | 5.3 | Th1 |
| LHFPL5 | 1487.8 | -0.731 | 0.167 | 0.0000113 | 0.00311 | N | 12.4 | Basophils |
| ARHGAP8 | 574.6 | -1.109 | 0.255 | 0.0000136 | 0.00353 | N | 26.6 | NK |
| LPL | 143.1 | -1.598 | 0.368 | 0.0000144 | 0.00358 | N | 19.4 | NC_mono |
| MT-ND2 | 171726.7 | -0.867 | 0.2 | 0.0000145 | 0.00358 | N | 3604.9 | PBMC |
| AS3MT | 284.9 | -1.354 | 0.318 | 0.0000205 | 0.00451 | N | 8.1 | Basophils |
| NDUFAB1P1 | 116.7 | -1.281 | 0.301 | 0.0000214 | 0.00451 | N | 17.6 | Basophils |
| NUS1 | 2704.6 | -0.551 | 0.129 | 0.0000202 | 0.00451 | N | 33.8 | pDC |
| MCC | 377 | -0.902 | 0.217 | 0.0000328 | 0.00606 | N | 19.3 | pDC |
| CEP44 | 2008.1 | -0.747 | 0.182 | 0.0000393 | 0.00688 | N | 30.6 | Basophils |
| DSC1 | 174.6 | -1.831 | 0.445 | 0.0000388 | 0.00688 | N | 36.8 | CD8_naive |
| C1QTNF3 | 409.2 | -0.729 | 0.18 | 0.0000513 | 0.00845 | N | 23.3 | Progenitor |
| SYNCRIP | 13662.6 | -0.476 | 0.119 | 0.0000614 | 0.00986 | N | 118.4 | Progenitor |
| MT-ND3 | 22435.6 | -0.71 | 0.179 | 0.0000692 | 0.0108 | N | 717.5 | B_Ex |
| PHEX | 200.7 | -1.053 | 0.266 | 0.0000734 | 0.0111 | N | 48.3 | pDC |
| SCAMP5 | 224.5 | -0.839 | 0.212 | 0.0000759 | 0.0114 | N | 353.9 | pDC |
| SPDYE2 | 20.1 | -2.908 | 0.738 | 0.0000806 | 0.0114 | N | 41.8 | TFH |
| TERF1 | 5972.3 | -0.644 | 0.164 | 0.0000861 | 0.0118 | N | 71.9 | NK |
| PRKD1 | 30.6 | -2.372 | 0.606 | 0.00009 | 0.0123 | N | 5 | B_NSM |
| CUX2 | 277.2 | -1.342 | 0.345 | 0.000102 | 0.0129 | N | 46.6 | pDC |
| CDKN3 | 266.7 | -0.806 | 0.208 | 0.000104 | 0.013 | N | 12.2 | Treg |
| SERPINF1 | 663.2 | -0.924 | 0.239 | 0.00011 | 0.0136 | N | 2551.9 | pDC |
| F2RL2 | 94.3 | -1.324 | 0.344 | 0.000118 | 0.0142 | N | 20 | NK |
| IBA57 | 2360.2 | -0.663 | 0.173 | 0.00013 | 0.0152 | N | 8 | Treg |
| CARD10 | 413.4 | -1.009 | 0.266 | 0.000145 | 0.0166 | N | 7.8 | C_mono |
| SRGAP2C | 4957.1 | -0.784 | 0.207 | 0.000152 | 0.017 | N | 20.5 | C_mono |
| PIP5KL1 | 124.2 | -1.037 | 0.274 | 0.000153 | 0.017 | N | 13.3 | pDC |
| CHPT1 | 2363.8 | -0.562 | 0.149 | 0.000161 | 0.0175 | N | 299.9 | Basophils |
| KRT5 | 80.7 | -1.901 | 0.508 | 0.000185 | 0.0195 | N | 233.6 | pDC |
| TSPAN19 | 225.1 | -0.817 | 0.22 | 0.000212 | 0.0214 | N | 10.3 | CD4_naive |
| LPAR1 | 952.6 | -0.989 | 0.269 | 0.000239 | 0.0238 | N | 33.8 | C_mono |
| TBL1XR1 | 10106.5 | -0.46 | 0.126 | 0.000252 | 0.0244 | N | 190.9 | pDC |
| ZNF92 | 2024.2 | -0.742 | 0.206 | 0.000307 | 0.0285 | N | 75 | B_SM |
| C4orf33 | 1082.8 | -0.535 | 0.149 | 0.000322 | 0.0297 | N | 52.7 | Basophils |
| VSNL1 | 191.8 | -1.051 | 0.293 | 0.000335 | 0.0308 | N | 5.7 | Basophils |
| POLR3F | 1075.2 | -0.497 | 0.139 | 0.000338 | 0.0309 | N | 34.7 | MAIT |
| ZNF519 | 2946.8 | -0.454 | 0.127 | 0.000349 | 0.0316 | N | 27.5 | CD8_naive |
| RAP1B | 37084.8 | -0.62 | 0.174 | 0.000374 | 0.0325 | N | 1262 | NK |
| SEH1L | 3878.9 | -0.693 | 0.195 | 0.000372 | 0.0325 | N | 66 | Progenitor |
| SLC22A16 | 185.4 | -1.212 | 0.341 | 0.000377 | 0.0327 | N | 16.7 | Progenitor |
| SRSF10P1 | 61.2 | -1.233 | 0.348 | 0.00039 | 0.0331 | N | 7.2 | Basophils |
| STT3B | 10152.6 | -0.448 | 0.127 | 0.000405 | 0.0338 | N | 124 | Progenitor |
| BZW2 | 1684.9 | -0.595 | 0.171 | 0.000493 | 0.0378 | N | 175.5 | Progenitor |
| MTATP6P1 | 24706.1 | -0.616 | 0.177 | 0.000487 | 0.0378 | N | 865.6 | B_NSM |
| PI4K2B | 1550.8 | -0.55 | 0.158 | 0.00049 | 0.0378 | N | 13.1 | VD2- |
| PSMG2 | 4274.8 | -0.641 | 0.184 | 0.000491 | 0.0378 | N | 131.4 | pDC |
| SRP9 | 3819.4 | -0.513 | 0.147 | 0.000492 | 0.0378 | N | 515.5 | CD8_EM |
| CYP4F29P | 64.3 | -2.483 | 0.713 | 0.0005 | 0.0382 | N | 140.5 | NK |
| AREG | 915.2 | -1.659 | 0.48 | 0.000539 | 0.0397 | N | 555.4 | Basophils |
| PLLP | 1129.3 | -0.595 | 0.172 | 0.000551 | 0.0397 | N | 18 | CD4_naive |
| SEPTIN7P14 | 158.9 | -1.065 | 0.308 | 0.000547 | 0.0397 | N | 187.3 | CD8_naive |
| FBXL2 | 551.9 | -0.583 | 0.169 | 0.000554 | 0.0398 | N | 10.5 | Progenitor |
| ZNF460 | 7132.3 | -0.467 | 0.135 | 0.000556 | 0.0398 | N | 38.5 | Th1 |
| TMEM87A | 3583.2 | -0.509 | 0.148 | 0.000577 | 0.041 | N | 187.4 | NK |
| SUSD4 | 364.9 | -0.867 | 0.252 | 0.000589 | 0.0417 | N | 28 | Th2 |
| BOC | 487.6 | -0.827 | 0.242 | 0.000629 | 0.0439 | N | 6.8 | B_Ex |
| SHCBP1 | 563.9 | -0.586 | 0.171 | 0.000636 | 0.0441 | N | 21.9 | Plasmablasts |
| ZMYM2 | 10308 | -0.478 | 0.14 | 0.00065 | 0.0444 | N | 79.6 | pDC |
| PDK1 | 7772.1 | -0.789 | 0.234 | 0.000744 | 0.0485 | N | 153.2 | CD4_naive |
| PLCH2 | 1547.1 | -0.833 | 0.247 | 0.000743 | 0.0485 | N | 14.9 | NK |
| RPL21P60 | 220.5 | -0.806 | 0.24 | 0.000771 | 0.0497 | N | 47 | TFH |
| MT-ATP6 | 107786.5 | -0.639 | 0.19 | 0.00078 | 0.04997 | N | 5083.6 | B_NSM |

